# Associations of Genetic Liability to Six Psychiatric Disorders With Cardiometabolic Diseases

**DOI:** 10.1101/2025.03.11.25323757

**Authors:** Jacob Bergstedt, Kadri Kõiv, Andreas Jangmo, Marit Haram, Piotr P. Jaholkowski, Jorien L. Treur, Isabell Brikell, Zheng Chang, Henrik Larsson, Patrik K. E. Magnusson, Andrew M. McIntosh, Cathryn M. Lewis, Brian K. Lee, Ida E. Sønderby, Yi Lu, Patrick F. Sullivan, Unnur A. Valdimarsdóttir, Ole Andreassen, Martin Tesli, Kelli Lehto, Fang Fang, the Anxiety Disorders Working Group of the Psychiatric Genomics Consortium, Estonian Biobank Research Team

## Abstract

**IMPORTANCE:** Individuals with psychiatric disorders have increased risk of cardiometabolic diseases (CMDs). Evaluating how psychiatric genetic liability relates to CMD may clarify mechanisms.

**OBJECTIVE:** Identify genetic overlap between psychiatric disorders and CMDs independent of cross-disorder pleiotropy, BMI, and smoking.

**DESIGN, SETTING, AND PARTICIPANTS:** Three Northern European cohorts (the Swedish Twin Registry, the Estonian Biobank, and the Norwegian Mother, Father and Child Cohort Study [MoBa]) totaling 355,159 individuals. Associations with CMDs were estimated as adjusted odds ratios (AORs) from logistic models mutually adjusted for all psychiatric PRSs and in models additionally adjusting for body mass index (BMI) and smoking. Cohort-specific AORs were pooled by inverse-variance weighting.

**MAIN OUTCOMES AND MEASURES:** Exposures were PRSs for attention-deficit/hyperactivity disorder (ADHD), major depressive disorder (MDD), anxiety disorder, posttraumatic stress disorder (PTSD), bipolar disorder, and schizophrenia. Outcomes were diagnoses of CMDs (hyperlipidemia, obesity, type 2 diabetes, hypertensive diseases, arteriosclerosis, ischemic heart disease, heart failure, thromboembolic disease, cerebrovascular disease, and arrhythmias), ascertained from electronic health records.

**RESULTS:** The MDD PRS was associated with increased risk of all CMDs across analyses (AORs ranged from 1.13 [95% CI, 1.10-1.15] for heart failure to 1.02 [95% CI, 1.00-1.05] for arrhythmias). The ADHD PRS was associated with increased risk of all CMDs (AOR ranged from 1.11 [95% CI, 1.09-1.12] for obesity to 1.02 [95% CI, 1.01-1.03] for hyperlipidemia), however associations where attenuated when adjusting for BMI and smoking (lifestyle adjusted AOR for obesity: 1.03 [95% CI, 1.02-1.05]). When not mutually adjusting for all psychiatric PRSs, anxiety disorder and PTSD PRSs were associated with all CMDs; these associations diminished after adjustment. The bipolar and schizophrenia PRSs were inversely associated with most CMDs (AOR for schizophrenia PRS and obesity, 0.93 [95% CI, 0.92-0.94]).

**CONCLUSIONS AND RELEVANCE:** Associations between psychiatric PRSs and CMDs diverged: ADHD, MDD, anxiety disorder, and PTSD PRSs were positively associated with CMDs, whereas bipolar and schizophrenia PRSs were inversely associated. Genetic liability to MDD showed robust associations with CMDs independent of cross-disorder pleiotropy, BMI, and smoking status, whereas associations between the ADHD PRS and CMDs were largely attenuated after adjustment for BMI and smoking.

**Key Points:** *Question:* Is genetic liability to specific psychiatric disorders associated with an increased risk of clinically diagnosed cardiometabolic diseases even after adjusting for cross-disorder psychiatric pleiotropy, body mass index, and smoking status?

*Findings:* In this study of cohorts in Sweden, Estonia, and Norway, genetic liability to major depressive disorder (MDD) showed stronger associations with cardiometabolic disease (CMD) compared to other psychiatric disorders. Genetic liability to anxiety disorder and PTSD showed associations with CMD that were attenuated when adjusting for cross-disorder psychiatric pleiotropy. Genetic liability to ADHD showed strong associations with obesity and type 2 diabetes that were strongly attenuated when adjusting for body mass index, and smoking status. Genetic liability to schizophrenia showed inverse associations with CMD.

*Meaning:* These findings suggest that MDD has a distinctive genetic relationship with CMD compared to other psychiatric disorders and that the well-established phenotypic association between schizophrenia and CMD is not related to genetic factors.

## Introduction

Cardiometabolic diseases (CMDs) reduce life expectancy in individuals with psychiatric disorders compared to the general population^1–3^. Studies using electronic health records have consistently shown that most psychiatric diagnoses are linked to CMD risk.^4,5^. Recent genome-wide association studies (GWASs) of psychiatric disorders have achieved sufficient statistical power to capture most common variant genetic risk^6,7^, which has enabled the construction of polygenic risk scores (PRSs; sums of risk alleles weighted by GWAS effect sizes) that capture individual-level genetic susceptibility^8^. Because PRSs are determined at birth, their associations with CMDs are immune to reverse causation and less prone to confounding than associations based on clinical diagnoses. PRS approaches have been employed to show that genetic liability to major depressive disorder (MDD)^9,10^, schizophrenia^9,10^, ADHD^11–13^, and post-traumatic stress disorder (PTSD)^14^ are associated with increased risk of CMDs. However, as psychiatric disorders exhibit substantial pleiotropy^15^, associations for a specific psychiatric PRS might reflect shared genetic background with other psychiatric disorders or lifestyle factors. Few studies have comprehensively mapped associations of genetic liability to psychiatric disorders with CMDs, while adjusting for other, genetically correlated, psychiatric PRSs or lifestyle factors.

We analyzed 355,159 genotyped participants from the population-based Swedish Twin Registry (STR), the volunteer-based Estonian Biobank (EstBB), and the population-based Norwegian Mother, Father, and Child Cohort Study (MoBa) to assess associations of PRSs for six psychiatric disorders (ADHD, MDD, anxiety disorder, PTSD, bipolar disorder, and schizophrenia) with clinical diagnoses of 10 CMDs. We estimated independent associations by mutually adjusting for all psychiatric PRSs and evaluated lifestyle influences through additional adjustment for body mass index (BMI) and smoking. Our primary objective was to identify genetic overlap between psychiatric disorders and CMDs independent of cross-disorder pleiotropy, BMI, and smoking, potentially reflecting shared pathophysiological pathways.

## Methods

### Study participants

### Swedish Twin Registry

The Screening Across the Lifespan Twin (SALT) cohort of the STR is a population-based study of all Swedish twins born between 1911 and 1958 conducted 1998–2002 (response rate of 70%^16^). Two studies nested in SALT were later conducted with the aim of genotyping the individuals. In the TwinGene study conducted 2004–2008, 22,000 SALT participants born in 1943 or earlier were invited to donate blood (response rate of 56%)^17^. In the SALTY study conducted 2008–2010, 24,914 SALT participants born 1943–1958 were invited to provide a saliva sample (response rate of 47%^18^). After quality control preprocessing, the STR sample comprised 17,378 genotyped individuals (Table 1).

**Table 1.** Description of the Swedish Twin Registry, Estonian Biobank, and MoBa cohorts. Life-time diagnoses until the end of follow-up are described. Records for hyperlipidemia were not available for the STR cohort. Records of obesity, cerebrovascular disease, thromboembolic disease, arrhythmias, and arteriosclerosis were not available for the MoBa cohort. ADHD, attention deficit/hyperactivity disorder; EstBB, Estonian Biobank; IQR, interquartile range; MDD, major depressive disorder; STR, Swedish Twin Registry.

|  |  | **STR** | **EstBB** | **MoBa** |
| --- | --- | --- | --- | --- |
| **Sample size** |  | 17,378 | 208,384 | 129,955 |
| **Age at end of follow-up, median (IQR), y** |  | 71 (65-77) | 51 (39-65) | 48 (41-55) |
| **Registers assessed** |  | In/out-patient care,<br>Cause-of-death | Primary care,<br>In/out-patient care,<br>Cause-of-death | In/out-patient<br>care |
| **Start of follow-up, y** |  | 1964 | 2004 | 2007 |
| **Start of follow-up with full coverage, y** |  | In-patient care (1987),<br>Out-patient care (2001),<br>cause-of-death (2004) | 2004 | 2007 |
| **End of follow-up** |  | 2016-12-31 | 2024-12-31 | 2022-12-31 |
| **Sex, n (%)** | **Female** | 9,046 (52.1) | 136,318 (65.4) | 77,064 (59.3) |
|  | **Male** | 8,332 (47.9) | 72,066 (34.6) | 52,891 (40.7) |
| **Psychiatric disorders, n (%)** | **ADHD** | 11 (0.1) | 3,282 (1.57) | 2,360 (1.8) |
|  | **MDD** | 862 (5.0) | 61,755 (29.6) | 11,043 (8.5) |
|  | **Anxiety disorders** | 505 (2.9) | 51,061 (24.5) | 8,325 (6.4) |
|  | **Stress-related disorders** | 350 (2.0) | 24,877 (11.9) | 10,018 (7.7) |
|  | **Bipolar disorder** | 147 (0.80) | 1,624 (0.8) | 1,420 (1.1) |
|  | **Schizophrenia** | 26 (0.1) | 1,084 (0.5) | 73 (0.05) |
| **Metabolic disorders, n (%)** | **Hyperlipidemia** | NA | 72,975 (35.0) | 2,268 (1.7) |
|  | **Type 2 diabetes** | 1,568 (9.0) | 18,528 (8.9) | 2,526 (1.9) |
|  | **Obesity** | 390 (2.2) | 38,355 (18.4) | NA |
| **Hypertensive diseases, n (%)** | **Hypertensive diseases** | 5,437 (31.3) | 81,040 (38.9) | 5,971 (4.6) |
| **CVDs, n (%)** | **Ischemic heart disease** | 2,537 (14.6) | 31,899 (15.3) | 2,248 (1.7) |
|  | **Heart failure** | 1,240 (7.1) | 21,149 (10.2) | 541 (0.4) |
|  | **Cerebrovascular disease** | 1,547 (8.9) | 19,630 (9.4) | NA |
|  | **Thromboembolic disease** | 967 (5.6) | 15,441 (7.4) | NA |
|  | **Arrhythmias** | 2,351 (13.5) | 16,261 (7.8) | NA |
|  | **Arteriosclerosis** | 822 (4.7) | 12,904 (6.2) | NA |

The use of the STR cohort in the present study was approved by the Swedish Ethical Review Authority (registration numbers 2021-03197, 2021-02994, 2021-02994).

### Estonian Biobank

The EstBB is a volunteer-based biobank with a sample size of 210,000 participants, comprising around 20% of the adult population of Estonia^19–21^ linked to primary and specialist care records.

The activities of the EstBB are regulated by the Human Genes Research Act, which was adopted in 2000 specifically for the operations of the EstBB. Individual level data analysis in the EstBB was carried out under ethical approval no. 1.1-12/624 and its extensions from the Estonian Committee on Bioethics and Human Research (Estonian Ministry of Social Affairs), using data according to release application no. 6-7/GI/16153 from the Estonian Biobank.

### MoBa

The MoBa study is conducted by the Norwegian Institute of Public Health. All pregnant women able to read Norwegian were eligible to participate in the study. Postal invitations were sent in advance of the first routine ultrasound examination in the 17^th^ week of pregnancy. Fathers were also invited to participate.

Participants were recruited 1999–2008 from all over Norway. The mothers consented to participation in 41% of the pregnancies. The cohort includes 114,500 individuals^22^. The establishment of MoBa and initial data collection was based on a license from the Norwegian Data Protection Agency and approval from The Regional committees for Medical and Health Research Ethics. The MoBa cohort is currently regulated by the Norwegian Health Registry Act. The use of MoBa cohort in the present study was approved by The Regional Committees for Medical and Health Research Ethics (REK 2016/1226).

### PRS profiling

We leveraged the most recent GWASs of ADHD^23^, MDD^6^, anxiety disorder^24^, PTSD^25^, bipolar disorder^26^, and schizophrenia^7^ to compute PRSs (eTable 1). Details on the genotyping in STR, EstBB, and MoBa are given in the Supplement (eMethods). If the discovery GWAS included samples from our study cohorts, we used leave-cohort-out GWAS summary statistics to compute PRSs. In EstBB, we removed 1,383 individuals (234 cases) that were included in a seminal schizophrenia GWAS study that formed the basis for subsequent studies^27^. We computed PRSs using the SBayesR framework^28^. We used a map of linkage disequilibrium estimated from 2,865,810 SNPs in 50,000 UK Biobank samples using a shrinkage estimator^28^. To reduce the contributions of horizontal pleiotropy, we additionally computed PRSs based solely on genome-wide significant variants using clumping and thresholding (P<5×10^−8^) implemented in PRSice-2^29^ (eTable 2). PRSs were standardized to have zero mean and a standard deviation of one.

### Outcome Measures

We ascertained psychiatric disorders and CMDs using electronic health records. We considered both main and secondary diagnoses. One diagnosis was sufficient to be identified as a case. To validate PRSs, we identified clinical diagnosis of ADHD, MDD, anxiety disorder, stress-related disorder, bipolar disorder, and schizophrenia (Table 1; eTable 3). Main outcomes were CMDs, including metabolic diseases (hyperlipidemia, type 2 diabetes [T2D], and obesity), hypertensive diseases, and cardiovascular diseases (CVDs; arteriosclerosis, ischemic heart diseases, heart failure, thromboembolic disease, cerebrovascular diseases, and arrhythmias; Table 1, eTable 4).

In STR, outcomes were identified from International Classification of Diseases (ICD)-8-10 codes in the Swedish National Patient Register (inpatient care nationwide since 1987 and specialist outpatient care with >80% national coverage since 2001) and from the Cause of Death Register, with follow-up through 2016. ADHD was excluded because of low prevalence (Table 1). Hyperlipidemia data were unavailable.

In the EstBB cohort, we used ICD-10 codes recorded in primary care, inpatient and outpatient care (Estonian Health Insurance Fund treatment bills, E-Health epicrises, and medical records from North Estonia Medical Centre and Tartu University Hospital), as well as the Cause of Death Register (since 2004), with follow-up through 2024.

In MoBa, we used ICD-10 codes from the Norwegian Patient Registry^30^ (all specialist health service visits nationwide since 2008) with follow-up through 2022. Data on obesity, cerebrovascular disease, thromboembolic disease, arrhythmias, and arteriosclerosis were not available for the MoBa cohort.

### Covariates

To assess the contribution of lifestyle factors, we adjusted for BMI and smoking. In the STR, BMI was computed from weight and height assessed in the SALT questionnaire (given 1998-2002). Smoking was also assessed in the SALT questionnaire and was categorized as: never (0), former (1), and current (2).

In the EstBB, BMI and smoking were assessed using the baseline questionnaire. BMI was computed from weight and height. Pregnant individuals at that time, extreme BMI outliers (values more than ±5 standard deviations from the mean), and BMI recorded before the age of 18 years were excluded. Smoking was categorized as: never (0), former (1), and current (2).

In the MoBa cohort, BMI and smoking status prior to pregnancy was assessed in baseline questionnaire that participating women answered at week 15 of pregnancy. The women reported on both their own and their partners BMI and smoking status. Since virtually no women smoked during pregnancy, smoking status in MoBa was assessed using the binary variable with categories: never (0), and former (1).

### Statistical Modeling

We conducted logistic regression analysis to estimate odds ratios (ORs) for the associations of psychiatric PRSs with psychiatric disorders (to validate PRSs) and CMDs. First, we performed a crude analysis where models were adjusted for the first ten genetic principal components (PCs), sex, and birth-year (modelled using a three degree-of-freedom natural spline term). To estimate adjusted ORs (AORs), we then conducted a mutually adjusted analysis additionally adjusted for all six PRSs, addressing pleiotropic effects across the six psychiatric disorders. In the STR cohort, we modelled sample correlation due to twinship using a sandwich estimator combined with generalized estimating equations (GEE). Pooled results were computed using inverse variance weighting of estimates from the three cohorts. Finally, we fitted models additionally adjusted for BMI and smoking status, to examine the potential influence of lifestyle factors on the studied associations. Proportion of variance explained on the liability scale, the log-liability scale, and according to Nagelkerkes measure^31^ were estimated in models including the PRS as the only predictor. We consider an association statistically significant if *P*<0.05.

## Results

### Association of psychiatric PRSs with psychiatric diagnoses

Psychiatric PRSs showed liability-scale R^2^ explained in psychiatric clinical diagnoses consistent with the original GWAS publications (liability-scale R^2^ ranged from 1–10%; eTable 5). In the crude analysis, all PRSs showed positive associations with all psychiatric disorders (eFigure 1), except that the ADHD PRS showed no association with schizophrenia. Cross-disorder associations were attenuated in the analysis with mutual adjustment of all psychiatric PRSs, indicating contributions of pleiotropy (eFigure 1). The strongest associations with diagnosis of the respective disorders were found for the schizophrenia and bipolar disorder PRSs (AORs, 1.77 and 1.34; *P*<1.66×10^−47^; eFigure 1; eTable 6). Estimates from individual cohorts showed similar patterns (eFigure 2).

### Association of psychiatric PRSs with CMD diagnoses

In the crude analysis, ADHD, MDD, anxiety disorder, and PTSD PRSs were associated with increased risk of all CMDs. The strongest associations were found for the ADHD and MDD PRSs with obesity (OR=1.16 for both PRSs, *P*<3.31×10^−59^; Figure 1; eTable 7) and for the MDD PRSs with heart failure (OR=1.16; *P*=8.34×10^−65^; Figure 1; eTable 7). Individual cohorts showed similar results, although the associations were consistently weaker in the EstBB cohort (eFigure 3).

**Figure 1.**
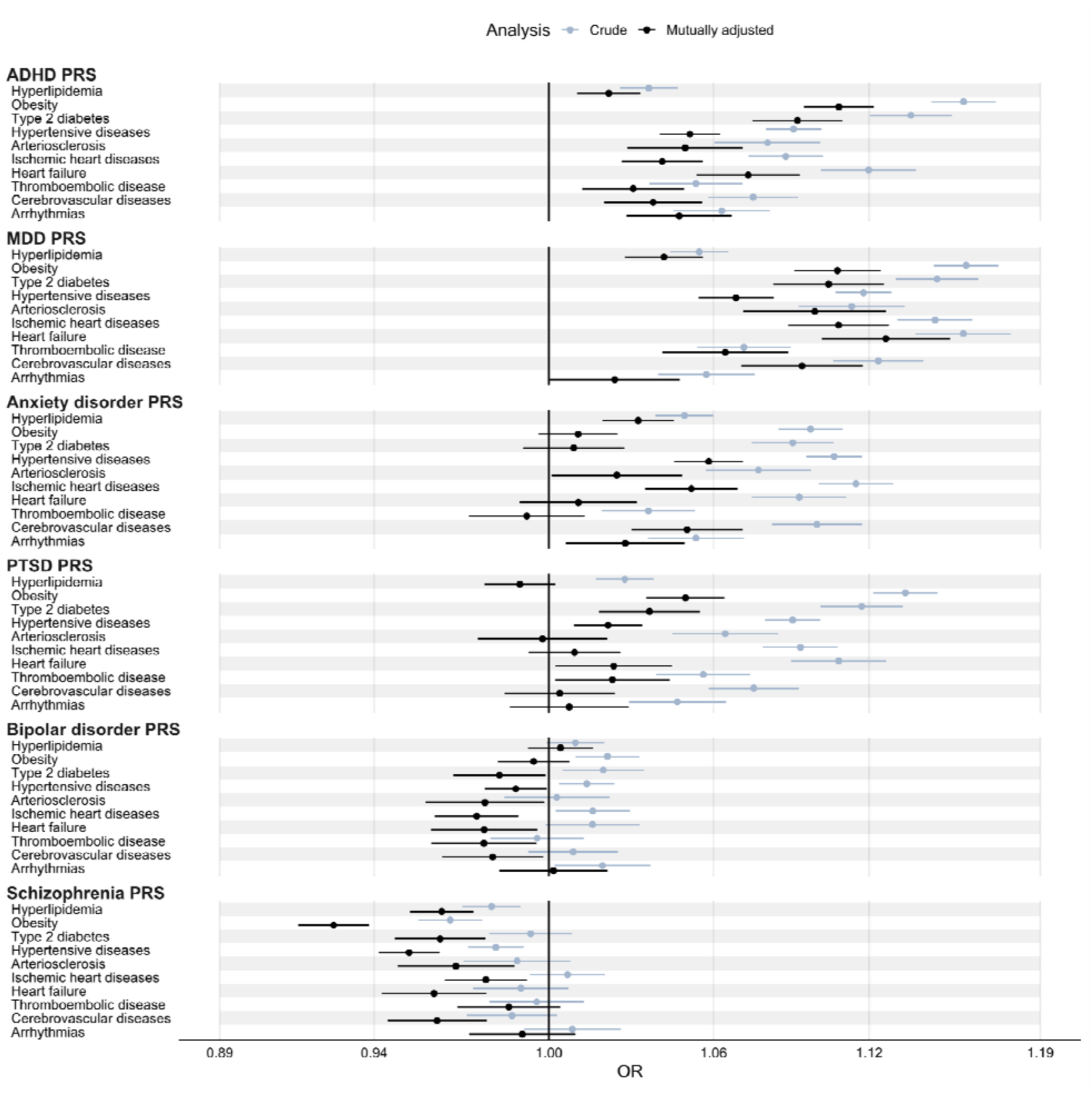
Associations of psychiatric PRSs with cardiometabolic diseases. In each cohort, ORs were computed from logistic regression models with the PRS as the exposure variable and any recorded clinica diagnosis of the cardiometabolic disease as the outcome variable. The crude model adjusted for the first 10 principal components, sex, and birth-year. The mutually adjusted model additionally adjusted for all other psychiatric PRSs. All ORs were computed from cohort-specific ORs using inverse variance weighting. All ORs correspond to the multiplicative change in odds per +1 standard deviation in PRS. ADHD, attention deficit/hyperactivity disorder; AOR, adjusted odds ratio; CMD, cardiometabolic disease; MDD, major depressive disorder; OR, odds ratio; PTSD, post-traumatic stress disorder; PRS, polygenic risk score.

The ADHD PRS showed statistically significant associations with all CMDs in the analysis mutually adjusted for all PRSs. The strongest associations were found with obesity and T2D (AORs, 1.11 and 1.09; *P*<1.81×10^−27^; Figure 1; eTable 7). The ADHD PRS based on genome-wide significant variants showed statistically significant associations for obesity, T2D, heart failure, and thromboembolic disease (eFigures 4-5).

The MDD PRS was associated with increased risk of all CMDs (except thromboembolic disease) across analyses (i.e., in models mutually adjusted for all PRSs and for the PRS based on genome-wide significant variants). It showed stronger associations with CVDs compared to the other psychiatric PRSs (nonoverlapping CIs indicated higher AORs for arteriosclerosis, ischemic heart disease, and heart failure, compared to all other PRSs; Figure 1; eFigures 4-5; eTable 7). The strongest association was for heart failure (AOR, 1.13; *P*=9.17×10^−25^; Figure 1; eTable 7).

The anxiety disorder PRS was associated with hyperlipidemia, hypertensive diseases, ischemic heart diseases, and cerebrovascular disease across analyses (Figure1; eTable 7; eFigures 4-5). The strongest association was for hypertensive disease (AOR, 1.06; *P*=9.60×10^−20^; Figure1; eTable 7).

For the PTSD PRS, associations with hyperlipidemia, arteriosclerosis, ischemic heart diseases, cerebrovascular disease, and arrhythmias were no longer statistically significant in the mutually adjusted analysis. It was associated with obesity, T2D, and hypertensive disease across analyses. The strongest AOR was for found for obesity (AOR, 1.05; *P*=8.46×10^−12^; Figure 1; eFigures 4-5; eTable 7).

The bipolar disorder PRS showed weak but statistically significant associations with obesity, T2D, hypertensive disease, ischemic heart diseases, and arrhythmias in the crude analysis (ORs<1.02; *P*>3.34×10^−4^; Figure 1; eTable 7). In contrast, it showed inverse associations with all CMDs except hyperlipidemia and arrhythmias in the mutually adjusted analysis (Figure 1). The strongest inverse association was found for ischemic heart diseases (AOR=0.97; P=7.06×10^−4^; Figure 1; eTable 7).

In the crude analysis, the schizophrenia PRS showed statistically significant inverse associations with hyperlipidemia, obesity, and hypertensive diseases (strongest for obesity: OR=0.97; *P*=1.67×10^−9^; Figure 1; eTable 7). After mutually adjustment for all other psychiatric PRSs, it showed statistically significant inverse associations with all CMDs except for thromboembolic diseases, and arrhythmias (strongest for obesity: AOR=0.93; *P*=5.11×10^−33^; Figure 1; eTable 7).

### Associations after adjustment for smoking status and BMI

The ADHD PRS showed attenuated AORs with CMDs after adjustment for BMI and smoking (lifestyle adjusted AORs<1.04; *P*>4.6×10^−5^; Figure 2; eTable 8). The largest attenuations were found for hyperlipidemia, obesity, T2D, and hypertensive diseases (82%, 69%, 60%, and 78% attenuation respectively; Figure 2).

**Figure 2.**
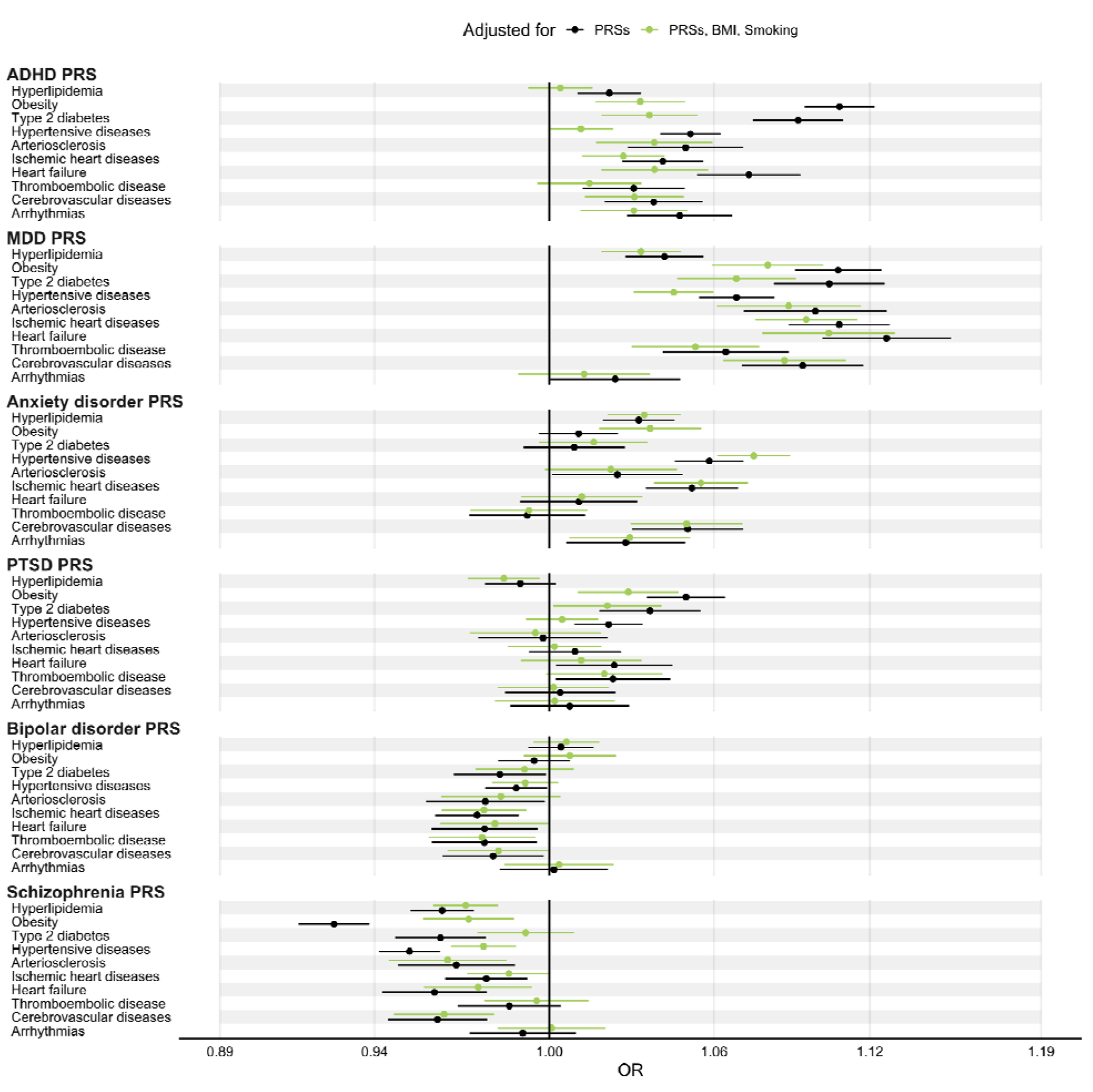
Associations of psychiatric PRSs with cardiometabolic diseases after adjustment for smoking status and BMI. In each cohort, ORs were computed from logistic regression models with the PRS as exposure variable and any recorded clinical diagnosis of the cardiometabolic disease as the outcome variable and all other psychiatric PRSs, sex, birth-year, and the 10 first genetic principal components as covariates. The “PRSs, BMI, Smoking” model additionally adjusted for BMI and smoking. All ORs were computed from cohort specific ORs by inverse variance weighting. All ORs correspond to the multiplicative change in odds per +1 standard deviation in PRS. ADHD, attention deficit/hyperactivity disorder; BMI, body mass index; CMD, cardiometabolic disease; MDD, major depressive disorder; OR, odds ratio; PTSD, post-traumatic stress disorder; PRS, polygenic risk score.

Associations for the MDD PRS were less affected than those for the ADHD PRS after adjustment for BMI and smoking. The largest attenuations were found for hypertensive diseases, T2D, and obesity (34%, 33%, and 24%). Cerebrovascular disease, arteriosclerosis, and ischemic heart disease showed limited attenuation (lifestyle adjusted AORs, 1.09, 1.09, and 1.10; reductions of 7.15%, 10.1%, and 11.4%; Figure 2; eTable 8). AORs for anxiety disorder, PTSD, and bipolar disorder PRSs showed also limited attenuation after adjustment for lifestyle factors.

In contrast, inverse associations of the schizophrenia PRS with obesity, T2D, and hypertensive diseases were attenuated after adjustment for BMI and smoking: ORs increased from 0.93 to 0.97, 0.96 to 0.99, and 0.95 to 0.98, respectively (reductions of 62.5%, 78.2%, and 52.8%).

## Discussion

The findings of this multinational study suggest that genetic liability to different psychiatric disorders show distinct patterns of association with CMDs. The MDD PRS showed the strongest associations with CMDs across analyses and cohorts. The ADHD PRS showed strong associations with obesity and T2D that were substantially attenuated when adjusting for BMI and smoking. In contrast, the schizophrenia PRS showed inverse associations with most CMDs across analyses and cohorts.

Psychiatric PRSs were positively associated with all psychiatric diagnoses in the crude analysis. After mutual adjustment for all psychiatric PRSs, most cross-disorder associations were substantially attenuated, consistent with extensive pleiotropy across disorders. Notably, the MDD PRS retained substantial cross-disorder associations even after mutual adjustment of all PRSs, suggesting that genetic overlap between other psychiatric PRSs and CMDs may be inflated by pleiotropy with MDD, highlighting the need for mutual adjustment.

The MDD PRS showed a distinctive association with increased CVD risk (arteriosclerosis, ischemic heart diseases, heart failure, thromboembolic disease, and cerebrovascular disease) relative to the other psychiatric PRSs. Combined with recent results based on time-to-event analysis in nationwide electronic health records^5^, genetic overlap estimated using GWAS summary data^32^, and two-sample Mendelian randomization^32^, our results add to increasingly robust support for a distinctive relationship between MDD and CVD.

The anxiety disorder PRS showed associations with increased risk of hyperlipidemia, hypertensive diseases, ischemic heart diseases, and cerebrovascular diseases across analyses providing robust evidence for a genetic link between anxiety disorder and CMDs, complementary to evidence based on register-based analyses^5^.

The PTSD PRS was associated with an increased risk of all CMDs in the crude analysis, confirming previous studies^14^. However, in the mutually adjusted analysis, associations with arteriosclerosis, ischemic heart diseases, cerebrovascular diseases, and arrhythmias disappeared, suggesting that these associations might be related to cross-disorder pleiotropy.

Schizophrenia PRSs were associated with a decreased risk of most CMDs, in contrast with previous reports^9,10,33^, but consistent with studies demonstrating a negative genetic correlation between schizophrenia and cardiometabolic risk factors^34–36^. The inverse associations became more pronounced after adjusting for other psychiatric PRSs, suggesting that pleiotropy with other psychiatric disorders might have inflated the crude estimates. A previous report found that individuals with untreated schizophrenia showed consistently lower BMI trajectories between 20–40 years of age compared to controls, whereas individuals with treated schizophrenia showed markedly increased BMI trajectories compared to controls, suggesting that our results are more in line with previous observations for untreated schizophrenia than treated schizophrenia^37^.

Psychiatric disorders are associated with altered BMI^37–39^ and increased likelihood of smoking^40–42^, which could contribute to the observed associations of psychiatric PRSs with CMDs. The ADHD PRS showed strong associations with metabolic and hypertensive diseases; however, after adjustment for BMI and smoking, these associations were largely attenuated, indicating that BMI and smoking play an important role in these relationships. By contrast, adjustment for BMI and smoking had little impact on the associations of MDD, PTSD, and anxiety disorder PRSs with CVDs, suggesting that BMI and smoking do not explain these genetic links. Notably, including BMI and smoking status as covariates attenuated the inverse associations observed for schizophrenia PRSs, consistent with previous reports of inverse associations between schizophrenia PRSs and BMI^35^.

### Limitations

Interpretation of the associations between psychiatric PRSs and CMDs remains challenging, as they may reflect pleiotropic effects. Psychiatric comorbidities are rarely excluded from the existing GWAS samples, potentially exacerbating psychiatric cross-disorder pleiotropy. Yet, consistent findings across analyses suggest that horizontal pleiotropy does not fully account for our observed associations.

All PRSs were derived from European-ancestry GWASs and applied to three Northern European cohorts, which may limit generalizability to other populations. The cohorts also differed in health-care context and data sources: STR included inpatient and outpatient data, MoBa inpatient data only, and EstBB primary care plus inpatient and outpatient data. However, results were broadly similar across cohorts, suggesting robustness to these differences.

BMI and smoking were assessed at a single time point, which may not capture their full effects; future work should consider longitudinal measures. Moreover, conditioning on post-exposure BMI or smoking may induce collider bias by opening paths between the PRS and factors that confound BMI/smoking– outcome relationships. Because such bias would likely affect all estimates similarly, it is unlikely to explain differences between PRSs.

We cannot exclude survival bias as a contributor to the inverse associations for schizophrenia and bipolar PRSs. These disorders are associated with premature mortality^2^; high PRS individuals maybe die of other causes before developing CMDs. Yet most psychiatric disorders are linked to premature mortality^2^, and late-onset diseases such as ischemic heart disease, which should be most sensitive to survival bias, showed among the weakest inverse associations. Together, these results suggest survival bias is unlikely to fully account for the inverse associations of schizophrenia and bipolar PRSs with CMDs.

Finally, differences in heritability, polygenicity, and GWAS sample size across psychiatric disorders complicate direct comparisons of association magnitudes across PRSs.

## Conclusions

Our multinational analyses provide robust evidence that MDD genetic liability is associated with increased risk of CMDs independent of cross-disorder psychiatric pleiotropy, BMI, and smoking. Associations between ADHD PRSs and CMDs were largely attenuated after adjustment for BMI and smoking. In contrast, schizophrenia PRSs showed inverse associations with several CMDs, suggesting that the well-documented phenotypic association between schizophrenia and CMDs is unlikely to be explained by shared common genetic liability.

## Data Availability

Data in this manuscript is available to authorized reserachers by formal application to the Swedish Twin Registry, the Estonian Biobank, and the MoBa study.

## Author Contributions

Drs Bergstedt, Kõiv, and Jangmo had full access to all data in the study and take responsibility for the integrity of the data and the accuracy of the data analysis. Drs Bergstedt, Kõiv, and Jangmo jointly contributed to this work. Drs Andreassen, Tesli, Lehto, and Fang jointly supervised this work.

*Concept and design:* Bergstedt, Kõiv, Jangmo, Andreassen, Tesli, Lehto, Fang

*Acquisition, analysis, or interpretation of data:* all authors

*Drafting of the manuscript:* Bergstedt, Kõiv, Jangmo, Andreassen, Tesli, Lehto, Fang

*Critical review of the manuscript for important intellectual content:* all authors

*Statistical analysis:* Bergstedt, Kõiv, Jangmo

*Obtained funding:* Lee, Sullivan, Valdimarsdóttir, Andreassen, Lehto, Fang

*Administrative, technical, or material support:* Bergstedt, Kõiv, Jangmo, Magnusson, Lu, Andreassen, Tesli, Lehto, Fang

*Supervision:* Andreassen, Tesli, Lehto, Fang

## Group information

The Anxiety Disorders Working Group of the Psychiatric Genomics Consortium and Estonian Biobank investigators are listed in the Supplement 2.

## Additional contributions

We thank the Estonian Biobank Research Team: Andres Metspalu, Lili Milani, Tõnu Esko, Reedik Mägi, Mait Metspalu, Mari Nelis and Georgi Hudjashov for data collection, genotyping,

