## Supplement 1 for "Associations of Genetic Liability to Six Psychiatric Disorders With Cardiometabolic Diseases"

### Supplement contents

**eTable 1.** GWAS sources used to construct psychiatric PRSs

**eTable 2**. Number of variants for PRSs derived using genome-wide significant variants

**eTable 3**. ICD codes for psychiatric disorders

**eTable 4**. ICD codes for cardiometabolic diseases

**eTable 5**. Measures of predictive performance (e.g., pseudo-R²) for psychiatric PRSs by cohort

**eTable 6**. Association estimates between psychiatric PRSs and clinical psychiatric diagnoses

**eTable 7**. Association estimates between psychiatric PRSs and cardiometabolic diseases

**eTable 8**. Association estimates between psychiatric PRSs and cardiometabolic diseases, adjusted for BMI and smoking

**eFigure 1**. Associations of psychiatric PRSs with clinical psychiatric diagnoses

**eFigure 2**. Associations of psychiatric PRSs with clinical psychiatric diagnoses by cohort

**eFigure 3**. Associations of psychiatric PRSs with cardiometabolic diseases by cohort

**eFigure 4**. Associations of psychiatric PRSs constructed from genome-wide significant loci with cardiometabolic diseases

**eFigure 5**. Associations of psychiatric PRSs constructed from genome-wide significant loci with cardiometabolic diseases by cohort

**eMethods**

### eTable 1. GWAS sources used to construct psychiatric PRSs

Case numbers from the GWASs including all cohorts (always excluding 23andMe). For PTSD, the case-control GWAS was used. Liability h^2^ is taken from the original publications.

ADHD, attention deficit/hyperactivity disorder; GWAS, genome-wide association study; MDD, major depressive disorder; PTSD, post-traumatic stress disorder.

| **Disorder** | **Discovery GWAS** | **PMID** | **N case** | **N controls** | **Liability h^2^** |
| --- | --- | --- | --- | --- | --- |
| **ADHD** | Demontis et al., 2023 | 36702997 | 38,691 | 275,986 | 22% |
| **MDD** | Adams et al., 2025 | 39814019 | 412,305 | 1,588,397 | 8.4% |
| **Anxiety disorder** | Strom et al., 2025 | 39006447 | 122,341 | 729,881 | 10.1% |
| **PTSD** | Nievergelt et al., 2024 | 38637617 | 140,767 | 1,109,073 | 5.3% |
| **Bipolar disorder** | O’Connell et al., 2025 | 39843750 | 59,282 | 781,022 | 11.3% |
| **Schizophrenia** | Trubetskoy et al., 2022 | 35396580 | 53,386 | 77,258 | 23% |

### eTable 2. Number of variants for PRSs derived using genome-wide significant variants

For STR, numbers are given for the TwinGene cohort (other STR cohorts were similar).

ADHD, attention deficit/hyperactivity disorder; EstBB, Estonian Biobank; MDD, major depressive disorder; PTSD, post-traumatic stress disorder; PRS, polygenic risk score; SNP, single nucleotide polymorphism; STR, Swedish Twin Registry.

| **Cohort** | **Disorder** | **Number of variants** |
| --- | --- | --- |
| **STR** | ADHD | 36 |
|  | MDD | 373 |
|  | Anxiety disorder | 75 |
|  | PTSD | 36 |
|  | Bipolar disorder | 103 |
|  | Schizophrenia | 280 |
| **EstBB** | ADHD | 37 |
|  | MDD | 333 |
|  | Anxiety disorder | 65 |
|  | PTSD | 24 |
|  | Bipolar disorder | 92 |
|  | Schizophrenia | 274 |
| **MoBa** | ADHD | 35 |
|  | MDD | 368 |
|  | Anxiety disorder | 63 |
|  | PTSD | 35 |
|  | Bipolar disorder | 95 |
|  | Schizophrenia | 349 |

### eTable 3. ICD codes for psychiatric disorders

ICD codes for psychiatric disorders. ICD 8-9 codes were only used in the Swedish Twin Registry.

ADHD, attention deficit/hyperactivity disorder; ICD, international classification of diseases; MDD, major depressive disorder.

| **Disorder** | **ICD-8** | **ICD-9** | **ICD-10** |
| --- | --- | --- | --- |
| **ADHD** |  | 314 | F90 |
| **MDD** | 296,2 300,4 298,0 | 296B 311 300E 298A | F32 F33 F341 F348 F349 F38 F39 |
| **Anxiety disorders** | 300,0 300,2 | 300A 300C | F40 F41 |
| **Stress-related disorders** | 307 | 308 309 | F43 |
| **Bipolar disorder** | 296,1 296,3 296,8 | 296A 296C 296D 296E 296F 296G 296H 296W 296X | F30 F31 |
| **Schizophrenia** | 295 | 295 | F20 |

### eTable 4. ICD codes for cardiometabolic diseases

ICD 8-9 codes were only used in the Swedish Twin Registry.

ICD, international classification of diseases

| **Disease** | **ICD-8** | **ICD-9** | **ICD-10** |
| --- | --- | --- | --- |
| **Hyperlipidemia** | 279 | 272 | E78 |
| **Obesity** | 277 | 278A 278B | E65 E66 |
| **Type 2 diabetes** | 250 | 250 | E11 |
| **Hypertensive diseases** | 400 401 402 403 404 | 401 402 403 404 405 | I1 |
| **Arteriosclerosis** | 440 441 442 443 444 | 440 441 442 443 444 | I70 I71 I72 I73 I74 |
| **Ischemic heart diseases** | 410 411 412 413 414 | 410 411 412 413 414 | I20 I21 I22 I23 I24 I251 I252 I255 I256 I258 I259 |
| **Heart failure** | 428 | 428 | I50 |
| **Arrhythmias** |  | 426A 426B 427A 427B 427D 427E 427F 427W | I441 I442 I46 I470 I471 I472 I48 I490 I495 I498 |
| **Thromboembolic disease** | 450 451 | 415B 451B | I26 I80 |
| **Cerebrovascular diseases** | 43 | 43 | I6 G45 |

### eTable 5: Measures of predictive performance (e.g., pseudo-R²) for psychiatric PRSs by cohort

Note that ADHD is missing for STR because of a lack of diagnosed cases in that cohort.

ADHD, attention deficit/hyperactivity disorder; EstBB, Estonian Biobank; MDD, major depressive disorder; PTSD, post-traumatic stress disorder; PRS, polygenic risk score; STR, Swedish Twin Registry.

| **Type** | **Value (%)** | **Cohort** | **Outcome** | **Exposure** |
| --- | --- | --- | --- | --- |
| R2 liability | 1.66 | STR | Anxiety disorder | Anxiety disorder PRS |
| R2 logit liability | 2.69 | STR | Anxiety disorder | Anxiety disorder PRS |
| Nagelkerke | 1.11 | STR | Anxiety disorder | Anxiety disorder PRS |
| R2 liability | 5.58 | STR | Bipolar disorder | Bipolar disorder PRS |
| R2 logit liability | 11.36 | STR | Bipolar disorder | Bipolar disorder PRS |
| Nagelkerke | 3.78 | STR | Bipolar disorder | Bipolar disorder PRS |
| R2 liability | 2.09 | STR | MDD | MDD PRS |
| R2 logit liability | 2.94 | STR | MDD | MDD PRS |
| Nagelkerke | 1.43 | STR | MDD | MDD PRS |
| R2 liability | 10.44 | STR | Schizophrenia | Schizophrenia PRS |
| R2 logit liability | 24.96 | STR | Schizophrenia | Schizophrenia PRS |
| Nagelkerke | 7.42 | STR | Schizophrenia | Schizophrenia PRS |
| R2 liability | 1.45 | STR | Stress-related disorders | PTSD PRS |
| R2 logit liability | 2.61 | STR | Stress-related disorders | PTSD PRS |
| Nagelkerke | 0.97 | STR | Stress-related disorders | PTSD PRS |
| R2 liability | 0.88 | EstBB | ADHD | ADHD PRS |
| R2 logit liability | 1.71 | EstBB | ADHD | ADHD PRS |
| Nagelkerke | 0.59 | EstBB | ADHD | ADHD PRS |
| R2 liability | 2.07 | EstBB | Anxiety disorder | Anxiety disorder PRS |
| R2 logit liability | 1.82 | EstBB | Anxiety disorder | Anxiety disorder PRS |
| Nagelkerke | 1.64 | EstBB | Anxiety disorder | Anxiety disorder PRS |
| R2 liability | 1.98 | EstBB | Bipolar disorder | Bipolar disorder PRS |
| R2 logit liability | 4.40 | EstBB | Bipolar disorder | Bipolar disorder PRS |
| Nagelkerke | 1.34 | EstBB | Bipolar disorder | Bipolar disorder PRS |
| R2 liability | 2.76 | EstBB | MDD | MDD PRS |
| R2 logit liability | 2.33 | EstBB | MDD | MDD PRS |
| Nagelkerke | 2.25 | EstBB | MDD | MDD PRS |
| R2 liability | 3.40 | EstBB | Schizophrenia | Schizophrenia PRS |
| R2 logit liability | 12.07 | EstBB | Schizophrenia | Schizophrenia PRS |
| Nagelkerke | 3.53 | EstBB | Schizophrenia | Schizophrenia PRS |
| R2 liability | 0.96 | EstBB | Stress-related disorders | PTSD PRS |
| R2 logit liability | 1.05 | EstBB | Stress-related disorders | PTSD PRS |
| Nagelkerke | 0.70 | EstBB | Stress-related disorders | PTSD PRS |
| R2 liability | 3.38 | MoBa | ADHD | ADHD PRS |
| **Type** | **Value (%)** | **Cohort** | **Outcome** | **Exposure** |
| R2 logit liability | 6.08 | MoBa | ADHD | ADHD PRS |
| Nagelkerke | 2.27 | MoBa | ADHD | ADHD PRS |
| R2 liability | 3.11 | MoBa | Anxiety disorder | Anxiety disorder PRS |
| R2 logit liability | 4.04 | MoBa | Anxiety disorder | Anxiety disorder PRS |
| Nagelkerke | 2.15 | MoBa | Anxiety disorder | Anxiety disorder PRS |
| R2 liability | 3.14 | MoBa | Bipolar disorder | Bipolar disorder PRS |
| R2 logit liability | 6.38 | MoBa | Bipolar disorder | Bipolar disorder PRS |
| Nagelkerke | 2.13 | MoBa | Bipolar disorder | Bipolar disorder PRS |
| R2 liability | 4.74 | MoBa | MDD | MDD PRS |
| R2 logit liability | 5.60 | MoBa | MDD | MDD PRS |
| Nagelkerke | 3.34 | MoBa | MDD | MDD PRS |
| R2 liability | 4.27 | MoBa | Schizophrenia | Schizophrenia PRS |
| R2 logit liability | 13.75 | MoBa | Schizophrenia | Schizophrenia PRS |
| Nagelkerke | 3.13 | MoBa | Schizophrenia | Schizophrenia PRS |
| R2 liability | 2.27 | MoBa | Stress-related disorders | PTSD PRS |
| R2 logit liability | 2.81 | MoBa | Stress-related disorders | PTSD PRS |
| Nagelkerke | 1.59 | MoBa | Stress-related disorders | PTSD PRS |

### eTable 6: Association estimates between psychiatric PRSs and clinical psychiatric diagnoses

ORs and P-values from crude and mutually adjusted models. In each cohort, ORs were computed using logistic regression models with the PRS as the exposure variable and any recorded clinical diagnosis of the psychiatric disorder as the outcome variable. The crude model adjusted for principal components, sex, and birth-year. Mutually adjusted models additionally adjusted for all psychiatric PRSs. ORs were computed from cohort-specific ORs by inverse variance weighting. ORs correspond to the multiplicative change in odds per +1 standard deviation in PRS

ADHD, attention deficit/hyperactivity disorder; ANX, anxiety disorder; AOR, adjusted odds ratio; BIP, bipolar disorder; MDD, major depressive disorder; mut. adj., mutually adjusted; OR, odds ratio; PTSD, post-traumatic stress disorder; PRS, polygenic risk score; SCZ, schizophrenia.

| **Exposure** | **Outcome** | **OR (Crude)** | **AOR (Mut. adj.)** | **P (Crude)** | **P (Mut. adj.)** |
| --- | --- | --- | --- | --- | --- |
| ADHD PRS | Hyperlipidemia | 1.04 [1.03, 1.05] | 1.02 [1.01, 1.03] | 1.42E-11 | 2.08E-04 |
| ADHD PRS | Obesity | 1.16 [1.14, 1.17] | 1.11 [1.09, 1.12] | 2.95E-141 | 3.31E-59 |
| ADHD PRS | Type 2 diabetes | 1.14 [1.12, 1.15] | 1.09 [1.07, 1.11] | 8.02E-67 | 1.81E-27 |
| ADHD PRS | Hypertensive diseases | 1.09 [1.08, 1.10] | 1.05 [1.04, 1.06] | 9.30E-67 | 7.62E-20 |
| ADHD PRS | Arteriosclerosis | 1.08 [1.06, 1.10] | 1.05 [1.03, 1.07] | 5.34E-16 | 3.73E-06 |
| ADHD PRS | Ischemic heart diseases | 1.09 [1.07, 1.10] | 1.04 [1.03, 1.06] | 1.16E-35 | 4.64E-08 |
| ADHD PRS | Heart failure | 1.12 [1.10, 1.14] | 1.07 [1.05, 1.09] | 9.33E-40 | 5.10E-14 |
| ADHD PRS | Thromboembolic disease | 1.05 [1.04, 1.07] | 1.03 [1.01, 1.05] | 7.53E-10 | 0.001 |
| ADHD PRS | Cerebrovascular diseases | 1.07 [1.06, 1.09] | 1.04 [1.02, 1.06] | 5.58E-19 | 3.19E-05 |
| ADHD PRS | Arrhythmias | 1.06 [1.05, 1.08] | 1.05 [1.03, 1.07] | 2.16E-12 | 1.19E-06 |
| MDD PRS | Hyperlipidemia | 1.05 [1.04, 1.07] | 1.04 [1.03, 1.06] | 5.29E-24 | 8.10E-09 |
| MDD PRS | Obesity | 1.16 [1.15, 1.17] | 1.11 [1.09, 1.12] | 7.42E-142 | 3.81E-39 |
| MDD PRS | Type 2 diabetes | 1.15 [1.13, 1.16] | 1.10 [1.08, 1.13] | 6.12E-76 | 4.76E-23 |
| MDD PRS | Hypertensive diseases | 1.12 [1.11, 1.13] | 1.07 [1.05, 1.08] | 6.44E-108 | 1.77E-22 |
| MDD PRS | Arteriosclerosis | 1.11 [1.09, 1.13] | 1.10 [1.07, 1.13] | 6.38E-29 | 2.93E-13 |
| MDD PRS | Ischemic heart diseases | 1.15 [1.13, 1.16] | 1.11 [1.09, 1.13] | 1.00E-90 | 1.92E-29 |
| MDD PRS | Heart failure | 1.16 [1.14, 1.18] | 1.13 [1.10, 1.15] | 8.34E-65 | 9.17E-25 |
| MDD PRS | Thromboembolic disease | 1.07 [1.05, 1.09] | 1.06 [1.04, 1.09] | 3.93E-16 | 4.14E-08 |
| MDD PRS | Cerebrovascular diseases | 1.12 [1.11, 1.14] | 1.09 [1.07, 1.12] | 4.25E-46 | 3.10E-16 |
| MDD PRS | Arrhythmias | 1.06 [1.04, 1.08] | 1.02 [1.00, 1.05] | 1.75E-10 | 0.047 |
| ANX PRS | Hyperlipidemia | 1.05 [1.04, 1.06] | 1.03 [1.02, 1.05] | 5.32E-20 | 1.08E-06 |
| ANX PRS | Obesity | 1.10 [1.08, 1.11] | 1.01 [1.00, 1.02] | 1.10E-57 | 0.149 |
| ANX PRS | Type 2 diabetes | 1.09 [1.07, 1.11] | 1.01 [0.99, 1.03] | 1.62E-31 | 0.340 |
| ANX PRS | Hypertensive diseases | 1.11 [1.10, 1.12] | 1.06 [1.05, 1.07] | 8.06E-90 | 9.60E-20 |
| ANX PRS | Arteriosclerosis | 1.08 [1.06, 1.10] | 1.02 [1.00, 1.05] | 7.00E-15 | 0.042 |
| ANX PRS | Ischemic heart diseases | 1.11 [1.10, 1.13] | 1.05 [1.03, 1.07] | 9.43E-59 | 1.51E-09 |
| ANX PRS | Heart failure | 1.09 [1.07, 1.11] | 1.01 [0.99, 1.03] | 2.98E-25 | 0.326 |
| ANX PRS | Thromboembolic disease | 1.04 [1.02, 1.05] | 0.99 [0.97, 1.01] | 3.07E-05 | 0.453 |
| ANX PRS | Cerebrovascular diseases | 1.10 [1.08, 1.12] | 1.05 [1.03, 1.07] | 1.50E-31 | 1.15E-06 |
| ANX PRS | Arrhythmias | 1.05 [1.04, 1.07] | 1.03 [1.01, 1.05] | 2.07E-09 | 0.012 |
| PTSD PRS | Hyperlipidemia | 1.03 [1.02, 1.04] | 0.99 [0.98, 1.00] | 3.13E-07 | 0.107 |
| PTSD PRS | Obesity | 1.13 [1.12, 1.15] | 1.05 [1.04, 1.06] | 1.26E-104 | 8.46E-12 |
| **Exposure** | **Outcome** | **OR (Crude)** | **AOR (Mut. adj.)** | **P (Crude)** | **P (Mut. adj.)** |
| PTSD PRS | Type 2 diabetes | 1.12 [1.10, 1.13] | 1.04 [1.02, 1.05] | 3.19E-50 | 9.53E-05 |
| PTSD PRS | Hypertensive diseases | 1.09 [1.08, 1.10] | 1.02 [1.01, 1.03] | 5.28E-66 | 7.07E-04 |
| PTSD PRS | Arteriosclerosis | 1.06 [1.04, 1.08] | 1.00 [0.98, 1.02] | 7.39E-11 | 0.846 |
| PTSD PRS | Ischemic heart diseases | 1.09 [1.08, 1.11] | 1.01 [0.99, 1.03] | 1.26E-39 | 0.276 |
| PTSD PRS | Heart failure | 1.11 [1.09, 1.13] | 1.02 [1.00, 1.04] | 9.18E-33 | 0.030 |
| PTSD PRS | Thromboembolic disease | 1.06 [1.04, 1.07] | 1.02 [1.00, 1.04] | 1.22E-10 | 0.030 |
| PTSD PRS | Cerebrovascular diseases | 1.08 [1.06, 1.09] | 1.00 [0.98, 1.02] | 6.66E-19 | 0.704 |
| PTSD PRS | Arrhythmias | 1.05 [1.03, 1.06] | 1.01 [0.99, 1.03] | 2.21E-07 | 0.504 |

### eTable 7: Association estimates between psychiatric PRSs and cardiometabolic diseases

ORs and P-values from crude and mutually adjusted models. In each cohort, ORs were computed using logistic regression models with the PRS as the exposure variable and any recorded clinical diagnosis of cardiometabolic disease as the outcome variable. The crude model adjusted for principal components, sex, and birth-year. Mutually adjusted models additionally adjusted for all psychiatric PRSs. ORs were computed from cohort-specific ORs by inverse variance weighting. ORs correspond to the multiplicative change in odds per +1 standard deviation in PRS

ADHD, attention deficit/hyperactivity disorder; ANX, anxiety disorder ; AOR, adjusted odds ratio; BIP, bipolar disorder; MDD, major depressive disorder; mut. adj., mutually adjusted; OR, odds ratio; PTSD, post-traumatic stress disorder; PRS, polygenic risk score; SCZ, schizophrenia.

| **Exposure** | **Outcome** | **OR (Crude)** | **AOR (Mut. adj.)** | **P (Crude)** | **P (Mut. adj.)** |
| --- | --- | --- | --- | --- | --- |
| ADHD PRS | Hyperlipidemia | 1.04 [1.03, 1.05] | 1.02 [1.01, 1.03] | 1.42E-11 | 2.08E-04 |
| ADHD PRS | Obesity | 1.16 [1.14, 1.17] | 1.11 [1.09, 1.12] | 2.95E-141 | 3.31E-59 |
| ADHD PRS | Type 2 diabetes | 1.14 [1.12, 1.15] | 1.09 [1.07, 1.11] | 8.02E-67 | 1.81E-27 |
| ADHD PRS | Hypertensive diseases | 1.09 [1.08, 1.10] | 1.05 [1.04, 1.06] | 9.30E-67 | 7.62E-20 |
| ADHD PRS | Arteriosclerosis | 1.08 [1.06, 1.10] | 1.05 [1.03, 1.07] | 5.34E-16 | 3.73E-06 |
| ADHD PRS | Ischemic heart diseases | 1.09 [1.07, 1.10] | 1.04 [1.03, 1.06] | 1.16E-35 | 4.64E-08 |
| ADHD PRS | Heart failure | 1.12 [1.10, 1.14] | 1.07 [1.05, 1.09] | 9.33E-40 | 5.10E-14 |
| ADHD PRS | Thromboembolic disease | 1.05 [1.04, 1.07] | 1.03 [1.01, 1.05] | 7.53E-10 | 0.001 |
| ADHD PRS | Cerebrovascular diseases | 1.07 [1.06, 1.09] | 1.04 [1.02, 1.06] | 5.58E-19 | 3.19E-05 |
| ADHD PRS | Arrhythmias | 1.06 [1.05, 1.08] | 1.05 [1.03, 1.07] | 2.16E-12 | 1.19E-06 |
| MDD PRS | Hyperlipidemia | 1.05 [1.04, 1.07] | 1.04 [1.03, 1.06] | 5.29E-24 | 8.10E-09 |
| MDD PRS | Obesity | 1.16 [1.15, 1.17] | 1.11 [1.09, 1.12] | 7.42E-142 | 3.81E-39 |
| MDD PRS | Type 2 diabetes | 1.15 [1.13, 1.16] | 1.10 [1.08, 1.13] | 6.12E-76 | 4.76E-23 |
| MDD PRS | Hypertensive diseases | 1.12 [1.11, 1.13] | 1.07 [1.05, 1.08] | 6.44E-108 | 1.77E-22 |
| MDD PRS | Arteriosclerosis | 1.11 [1.09, 1.13] | 1.10 [1.07, 1.13] | 6.38E-29 | 2.93E-13 |
| MDD PRS | Ischemic heart diseases | 1.15 [1.13, 1.16] | 1.11 [1.09, 1.13] | 1.00E-90 | 1.92E-29 |
| MDD PRS | Heart failure | 1.16 [1.14, 1.18] | 1.13 [1.10, 1.15] | 8.34E-65 | 9.17E-25 |
| MDD PRS | Thromboembolic disease | 1.07 [1.05, 1.09] | 1.06 [1.04, 1.09] | 3.93E-16 | 4.14E-08 |
| MDD PRS | Cerebrovascular diseases | 1.12 [1.11, 1.14] | 1.09 [1.07, 1.12] | 4.25E-46 | 3.10E-16 |
| MDD PRS | Arrhythmias | 1.06 [1.04, 1.08] | 1.02 [1.00, 1.05] | 1.75E-10 | 0.047 |
| ANX PRS | Hyperlipidemia | 1.05 [1.04, 1.06] | 1.03 [1.02, 1.05] | 5.32E-20 | 1.08E-06 |
| ANX PRS | Obesity | 1.10 [1.08, 1.11] | 1.01 [1.00, 1.02] | 1.10E-57 | 0.149 |
| ANX PRS | Type 2 diabetes | 1.09 [1.07, 1.11] | 1.01 [0.99, 1.03] | 1.62E-31 | 0.340 |
| ANX PRS | Hypertensive diseases | 1.11 [1.10, 1.12] | 1.06 [1.05, 1.07] | 8.06E-90 | 9.60E-20 |
| ANX PRS | Arteriosclerosis | 1.08 [1.06, 1.10] | 1.02 [1.00, 1.05] | 7.00E-15 | 0.042 |
| ANX PRS | Ischemic heart diseases | 1.11 [1.10, 1.13] | 1.05 [1.03, 1.07] | 9.43E-59 | 1.51E-09 |
| ANX PRS | Heart failure | 1.09 [1.07, 1.11] | 1.01 [0.99, 1.03] | 2.98E-25 | 0.326 |
| ANX PRS | Thromboembolic disease | 1.04 [1.02, 1.05] | 0.99 [0.97, 1.01] | 3.07E-05 | 0.453 |
| ANX PRS | Cerebrovascular diseases | 1.10 [1.08, 1.12] | 1.05 [1.03, 1.07] | 1.50E-31 | 1.15E-06 |
| ANX PRS | Arrhythmias | 1.05 [1.04, 1.07] | 1.03 [1.01, 1.05] | 2.07E-09 | 0.012 |
| PTSD PRS | Hyperlipidemia | 1.03 [1.02, 1.04] | 0.99 [0.98, 1.00] | 3.13E-07 | 0.107 |
| PTSD PRS | Obesity | 1.13 [1.12, 1.15] | 1.05 [1.04, 1.06] | 1.26E-104 | 8.46E-12 |
| **Exposure** | **Outcome** | **OR (Crude)** | **AOR (Mut. adj.)** | **P (Crude)** | **P (Mut. adj.)** |
| PTSD PRS | Type 2 diabetes | 1.12 [1.10, 1.13] | 1.04 [1.02, 1.05] | 3.19E-50 | 9.53E-05 |
| PTSD PRS | Hypertensive diseases | 1.09 [1.08, 1.10] | 1.02 [1.01, 1.03] | 5.28E-66 | 7.07E-04 |
| PTSD PRS | Arteriosclerosis | 1.06 [1.04, 1.08] | 1.00 [0.98, 1.02] | 7.39E-11 | 0.846 |
| PTSD PRS | Ischemic heart diseases | 1.09 [1.08, 1.11] | 1.01 [0.99, 1.03] | 1.26E-39 | 0.276 |
| PTSD PRS | Heart failure | 1.11 [1.09, 1.13] | 1.02 [1.00, 1.04] | 9.18E-33 | 0.030 |
| PTSD PRS | Thromboembolic disease | 1.06 [1.04, 1.07] | 1.02 [1.00, 1.04] | 1.22E-10 | 0.030 |
| PTSD PRS | Cerebrovascular diseases | 1.08 [1.06, 1.09] | 1.00 [0.98, 1.02] | 6.66E-19 | 0.704 |
| PTSD PRS | Arrhythmias | 1.05 [1.03, 1.06] | 1.01 [0.99, 1.03] | 2.21E-07 | 0.504 |
| BIP PRS | Hyperlipidemia | 1.01 [1.00, 1.02] | 1.00 [0.99, 1.02] | 0.074 | 0.488 |
| BIP PRS | Obesity | 1.02 [1.01, 1.03] | 0.99 [0.98, 1.01] | 3.34E-04 | 0.405 |
| BIP PRS | Type 2 diabetes | 1.02 [1.00, 1.03] | 0.98 [0.97, 1.00] | 0.009 | 0.035 |
| BIP PRS | Hypertensive diseases | 1.01 [1.00, 1.02] | 0.99 [0.98, 1.00] | 0.007 | 0.036 |
| BIP PRS | Arteriosclerosis | 1.00 [0.98, 1.02] | 0.98 [0.96, 1.00] | 0.771 | 0.034 |
| BIP PRS | Ischemic heart diseases | 1.02 [1.00, 1.03] | 0.97 [0.96, 0.99] | 0.021 | 7.06E-04 |
| BIP PRS | Heart failure | 1.02 [1.00, 1.03] | 0.98 [0.96, 1.00] | 0.070 | 0.017 |
| BIP PRS | Thromboembolic disease | 1.00 [0.98, 1.01] | 0.98 [0.96, 1.00] | 0.620 | 0.015 |
| BIP PRS | Cerebrovascular diseases | 1.01 [0.99, 1.02] | 0.98 [0.96, 1.00] | 0.290 | 0.029 |
| BIP PRS | Arrhythmias | 1.02 [1.00, 1.04] | 1.00 [0.98, 1.02] | 0.029 | 0.869 |
| SCZ PRS | Hyperlipidemia | 0.98 [0.97, 0.99] | 0.96 [0.95, 0.97] | 1.13E-04 | 4.69E-11 |
| SCZ PRS | Obesity | 0.97 [0.95, 0.98] | 0.93 [0.92, 0.94] | 1.67E-09 | 5.11E-33 |
| SCZ PRS | Type 2 diabetes | 0.99 [0.98, 1.01] | 0.96 [0.95, 0.98] | 0.384 | 2.42E-06 |
| SCZ PRS | Hypertensive diseases | 0.98 [0.97, 0.99] | 0.95 [0.94, 0.96] | 1.86E-04 | 2.65E-19 |
| SCZ PRS | Arteriosclerosis | 0.99 [0.97, 1.01] | 0.97 [0.95, 0.99] | 0.241 | 0.002 |
| SCZ PRS | Ischemic heart diseases | 1.01 [0.99, 1.02] | 0.98 [0.96, 0.99] | 0.331 | 0.003 |
| SCZ PRS | Heart failure | 0.99 [0.97, 1.01] | 0.96 [0.94, 0.98] | 0.249 | 1.65E-05 |
| SCZ PRS | Thromboembolic disease | 1.00 [0.98, 1.01] | 0.99 [0.97, 1.00] | 0.611 | 0.128 |
| SCZ PRS | Cerebrovascular diseases | 0.99 [0.97, 1.00] | 0.96 [0.94, 0.98] | 0.109 | 1.01E-05 |
| SCZ PRS | Arrhythmias | 1.01 [0.99, 1.03] | 0.99 [0.97, 1.01] | 0.344 | 0.325 |

### eTable 8: Association estimates between psychiatric PRSs and cardiometabolic diseases, adjusted for BMI and smoking

ORs and P-values from models adjusted for all psychiatric PRSs and models adjusted for all psychiatric PRSs, BMI and smoking. In each cohort, ORs were computed using logistic regression models with the PRS as the exposure variable and any recorded clinical diagnosis of the cardiometabolic disease as the as the outcome variable. The models additionally adjusted for principal components, sex, and birth-year. ORs were computed from cohort-specific ORs by inverse variance weighting. ORs correspond to the multiplicative change in odds per +1 standard deviation in PRS.

Adj. on PRSs, lifestyle, adjusted on PRSs and lifestyle; ADHD, attention deficit/hyperactivity disorder; ANX, anxiety disorder; BIP, bipolar disorder; MDD, major depressive disorder; Mut. adj., mutually adjusted; OR, odds ratio; PTSD, post-traumatic stress disorder; PRS, polygenic risk score; SCZ, schizophrenia.

| **Exposure** | **Outcome** | **AOR (Mut. adj.)** | **AOR (Adj. on PRSs, lifestyle)** | **P (Mut. adj.)** | **P (Adj. on PRSs, lifestyle)** |
| --- | --- | --- | --- | --- | --- |
| ADHD PRS | Hyperlipidemia | 1.02 [1.01, 1.03] | 1.00 [0.99, 1.02] | 2.08E-04 | 0.507 |
| ADHD PRS | Obesity | 1.11 [1.09, 1.12] | 1.03 [1.02, 1.05] | 3.31E-59 | 7.74E-05 |
| ADHD PRS | Type 2 diabetes | 1.09 [1.07, 1.11] | 1.04 [1.02, 1.05] | 1.81E-27 | 4.63E-05 |
| ADHD PRS | Hypertensive diseases | 1.05 [1.04, 1.06] | 1.01 [1.00, 1.02] | 7.62E-20 | 0.056 |
| ADHD PRS | Arteriosclerosis | 1.05 [1.03, 1.07] | 1.04 [1.02, 1.06] | 3.73E-06 | 4.29E-04 |
| ADHD PRS | Ischemic heart diseases | 1.04 [1.03, 1.06] | 1.03 [1.01, 1.04] | 4.64E-08 | 4.79E-04 |
| ADHD PRS | Heart failure | 1.07 [1.05, 1.09] | 1.04 [1.02, 1.06] | 5.10E-14 | 1.22E-04 |
| ADHD PRS | Thromboembolic disease | 1.03 [1.01, 1.05] | 1.01 [1.00, 1.03] | 0.001 | 0.132 |
| ADHD PRS | Cerebrovascular diseases | 1.04 [1.02, 1.06] | 1.03 [1.01, 1.05] | 3.19E-05 | 7.93E-04 |
| ADHD PRS | Arrhythmias | 1.05 [1.03, 1.07] | 1.03 [1.01, 1.05] | 1.19E-06 | 0.002 |
| MDD PRS | Hyperlipidemia | 1.04 [1.03, 1.06] | 1.03 [1.02, 1.05] | 8.10E-09 | 6.53E-06 |
| MDD PRS | Obesity | 1.11 [1.09, 1.12] | 1.08 [1.06, 1.10] | 3.81E-39 | 1.38E-14 |
| MDD PRS | Type 2 diabetes | 1.10 [1.08, 1.13] | 1.07 [1.05, 1.09] | 4.76E-23 | 6.67E-10 |
| MDD PRS | Hypertensive diseases | 1.07 [1.05, 1.08] | 1.04 [1.03, 1.06] | 1.77E-22 | 1.14E-09 |
| MDD PRS | Arteriosclerosis | 1.10 [1.07, 1.13] | 1.09 [1.06, 1.12] | 2.93E-13 | 7.86E-11 |
| MDD PRS | Ischemic heart diseases | 1.11 [1.09, 1.13] | 1.10 [1.08, 1.12] | 1.92E-29 | 9.39E-23 |
| MDD PRS | Heart failure | 1.13 [1.10, 1.15] | 1.10 [1.08, 1.13] | 9.17E-25 | 1.57E-16 |
| MDD PRS | Thromboembolic disease | 1.06 [1.04, 1.09] | 1.05 [1.03, 1.08] | 4.14E-08 | 7.70E-06 |
| MDD PRS | Cerebrovascular diseases | 1.09 [1.07, 1.12] | 1.09 [1.06, 1.11] | 3.10E-16 | 5.50E-14 |
| MDD PRS | Arrhythmias | 1.02 [1.00, 1.05] | 1.01 [0.99, 1.04] | 0.047 | 0.303 |
| ANX PRS | Hyperlipidemia | 1.03 [1.02, 1.05] | 1.03 [1.02, 1.05] | 1.08E-06 | 3.44E-07 |
| ANX PRS | Obesity | 1.01 [1.00, 1.02] | 1.04 [1.02, 1.06] | 0.149 | 1.04E-04 |
| ANX PRS | Type 2 diabetes | 1.01 [0.99, 1.03] | 1.02 [1.00, 1.04] | 0.340 | 0.111 |
| ANX PRS | Hypertensive diseases | 1.06 [1.05, 1.07] | 1.07 [1.06, 1.09] | 9.60E-20 | 8.94E-28 |
| ANX PRS | Arteriosclerosis | 1.02 [1.00, 1.05] | 1.02 [1.00, 1.05] | 0.042 | 0.068 |
| ANX PRS | Ischemic heart diseases | 1.05 [1.03, 1.07] | 1.06 [1.04, 1.07] | 1.51E-09 | 2.55E-10 |
| ANX PRS | Heart failure | 1.01 [0.99, 1.03] | 1.01 [0.99, 1.03] | 0.326 | 0.295 |
| ANX PRS | Thromboembolic disease | 0.99 [0.97, 1.01] | 0.99 [0.97, 1.01] | 0.453 | 0.495 |
| ANX PRS | Cerebrovascular diseases | 1.05 [1.03, 1.07] | 1.05 [1.03, 1.07] | 1.15E-06 | 1.69E-06 |
| ANX PRS | Arrhythmias | 1.03 [1.01, 1.05] | 1.03 [1.01, 1.05] | 0.012 | 0.009 |
| PTSD PRS | Hyperlipidemia | 0.99 [0.98, 1.00] | 0.98 [0.97, 1.00] | 0.107 | 0.013 |
| **Exposure** | **Outcome** | **AOR (Mut. adj.)** | **AOR (Adj. on PRSs, lifestyle)** | **P (Mut. adj.)** | **P (Adj. on PRSs, lifestyle)** |
| PTSD PRS | Obesity | 1.05 [1.04, 1.06] | 1.03 [1.01, 1.05] | 8.46E-12 | 0.002 |
| PTSD PRS | Type 2 diabetes | 1.04 [1.02, 1.05] | 1.02 [1.00, 1.04] | 9.53E-05 | 0.036 |
| PTSD PRS | Hypertensive diseases | 1.02 [1.01, 1.03] | 1.00 [0.99, 1.02] | 7.07E-04 | 0.491 |
| PTSD PRS | Arteriosclerosis | 1.00 [0.98, 1.02] | 1.00 [0.97, 1.02] | 0.846 | 0.674 |
| PTSD PRS | Ischemic heart diseases | 1.01 [0.99, 1.03] | 1.00 [0.99, 1.02] | 0.276 | 0.834 |
| PTSD PRS | Heart failure | 1.02 [1.00, 1.04] | 1.01 [0.99, 1.03] | 0.030 | 0.301 |
| PTSD PRS | Thromboembolic disease | 1.02 [1.00, 1.04] | 1.02 [1.00, 1.04] | 0.030 | 0.065 |
| PTSD PRS | Cerebrovascular diseases | 1.00 [0.98, 1.02] | 1.00 [0.98, 1.02] | 0.704 | 0.890 |
| PTSD PRS | Arrhythmias | 1.01 [0.99, 1.03] | 1.00 [0.98, 1.02] | 0.504 | 0.864 |
| BIP PRS | Hyperlipidemia | 1.00 [0.99, 1.02] | 1.01 [0.99, 1.02] | 0.488 | 0.313 |
| BIP PRS | Obesity | 0.99 [0.98, 1.01] | 1.01 [0.99, 1.02] | 0.405 | 0.388 |
| BIP PRS | Type 2 diabetes | 0.98 [0.97, 1.00] | 0.99 [0.97, 1.01] | 0.035 | 0.322 |
| BIP PRS | Hypertensive diseases | 0.99 [0.98, 1.00] | 0.99 [0.98, 1.00] | 0.036 | 0.153 |
| BIP PRS | Arteriosclerosis | 0.98 [0.96, 1.00] | 0.98 [0.96, 1.00] | 0.034 | 0.112 |
| BIP PRS | Ischemic heart diseases | 0.97 [0.96, 0.99] | 0.98 [0.96, 0.99] | 7.06E-04 | 0.003 |
| BIP PRS | Heart failure | 0.98 [0.96, 1.00] | 0.98 [0.96, 1.00] | 0.017 | 0.052 |
| BIP PRS | Thromboembolic disease | 0.98 [0.96, 1.00] | 0.98 [0.96, 1.00] | 0.015 | 0.013 |
| BIP PRS | Cerebrovascular diseases | 0.98 [0.96, 1.00] | 0.98 [0.96, 1.00] | 0.029 | 0.050 |
| BIP PRS | Arrhythmias | 1.00 [0.98, 1.02] | 1.00 [0.98, 1.02] | 0.869 | 0.733 |
| SCZ PRS | Hyperlipidemia | 0.96 [0.95, 0.97] | 0.97 [0.96, 0.98] | 4.69E-11 | 4.39E-07 |
| SCZ PRS | Obesity | 0.93 [0.92, 0.94] | 0.97 [0.96, 0.99] | 5.11E-33 | 4.65E-04 |
| SCZ PRS | Type 2 diabetes | 0.96 [0.95, 0.98] | 0.99 [0.97, 1.01] | 2.42E-06 | 0.336 |
| SCZ PRS | Hypertensive diseases | 0.95 [0.94, 0.96] | 0.98 [0.97, 0.99] | 2.65E-19 | 6.78E-05 |
| SCZ PRS | Arteriosclerosis | 0.97 [0.95, 0.99] | 0.96 [0.94, 0.98] | 0.002 | 7.22E-04 |
| SCZ PRS | Ischemic heart diseases | 0.98 [0.96, 0.99] | 0.99 [0.97, 1.00] | 0.003 | 0.057 |
| SCZ PRS | Heart failure | 0.96 [0.94, 0.98] | 0.98 [0.96, 0.99] | 1.65E-05 | 0.010 |
| SCZ PRS | Thromboembolic disease | 0.99 [0.97, 1.00] | 1.00 [0.98, 1.01] | 0.128 | 0.631 |
| SCZ PRS | Cerebrovascular diseases | 0.96 [0.94, 0.98] | 0.96 [0.95, 0.98] | 1.01E-05 | 3.64E-05 |
| SCZ PRS | Arrhythmias | 0.99 [0.97, 1.01] | 1.00 [0.98, 1.02] | 0.325 | 0.938 |

### eFigure 1. Associations of psychiatric PRSs with clinical psychiatric diagnoses

In each cohort, ORs were computed using logistic regression models with one psychiatric PRS as the exposure variable and any recorded clinical diagnosis of a psychiatric disorder as the outcome variable. The crude model adjusted for the first 10 principal components, sex, and birth-year. The adjusted model additionally adjusted for all other psychiatric PRSs. ORs were computed from cohort-specific ORs using inverse variance weighting. ORs correspond to the multiplicative change in odds per +1 standard deviation in PRS. ADHD, attention deficit/hyperactivity disorder; AOR, adjusted odds ratio; MDD, major depressive disorder; OR, odds ratio; PTSD, post-traumatic stress disorder; PRS, polygenic risk score.

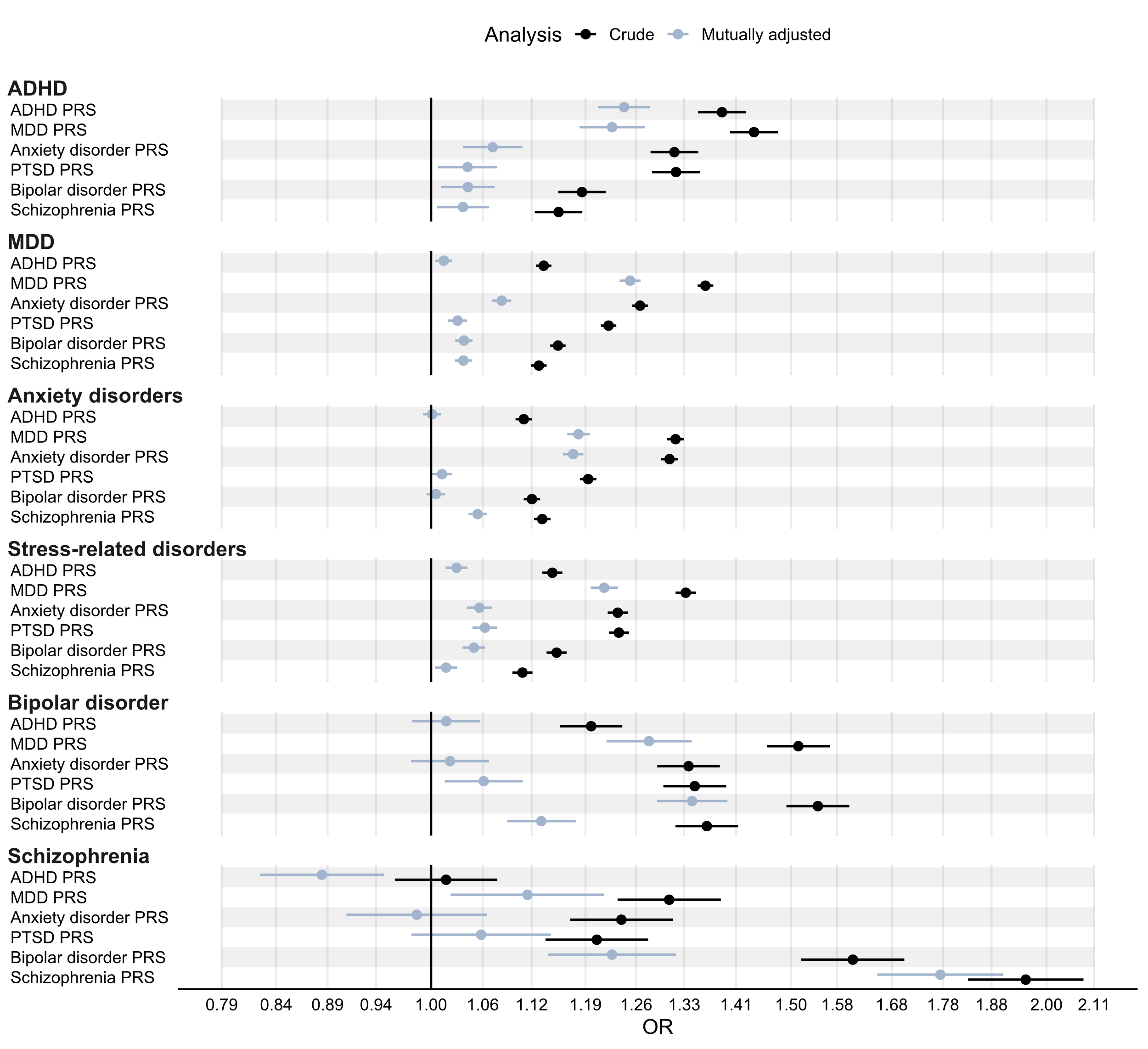

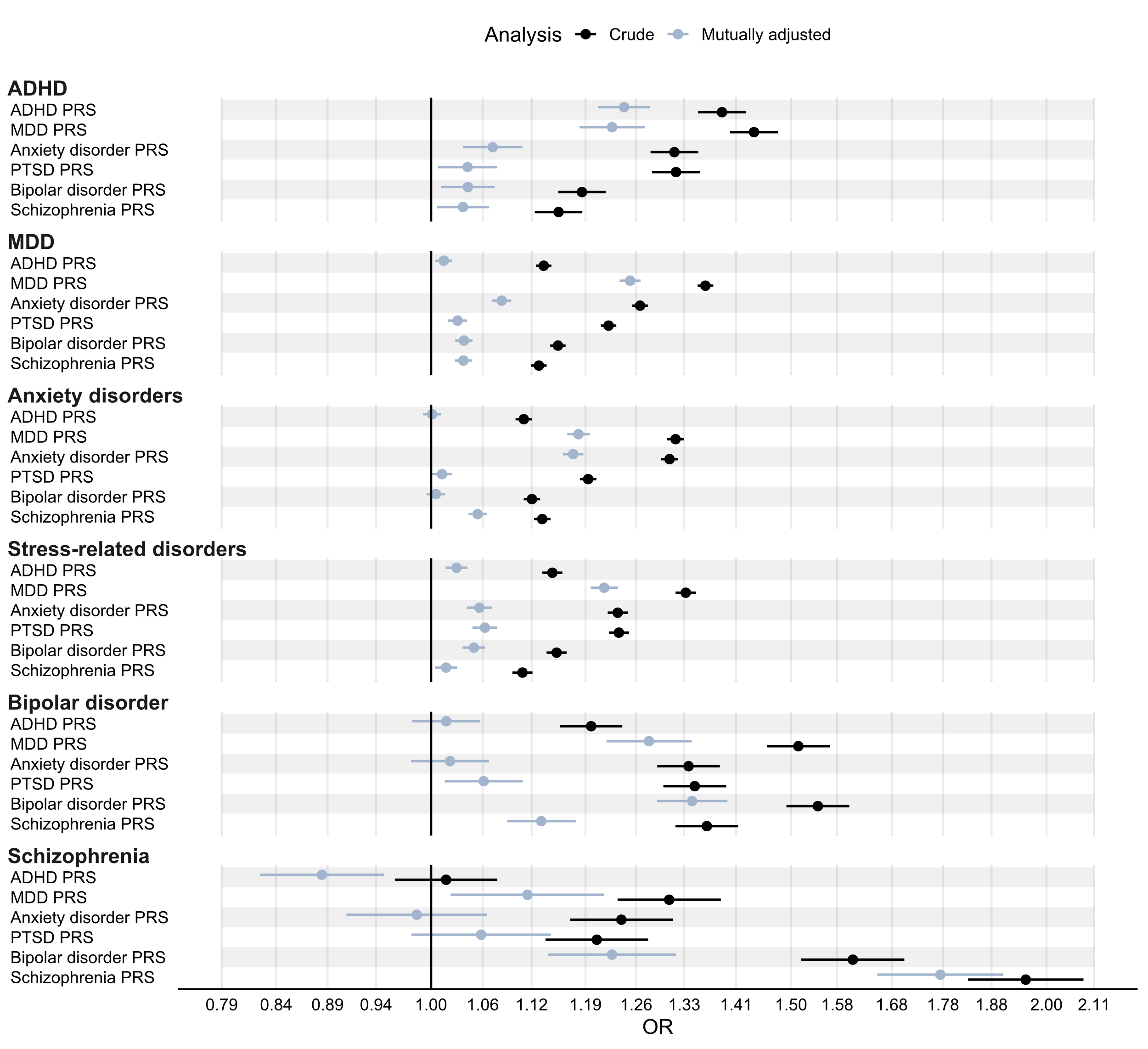

### eFigure 2. Associations of psychiatric PRSs with clinical psychiatric diagnoses by cohort

In each cohort, ORs were computed using logistic regression models with the PRS as exposure variable and any recorded clinical diagnosis of the psychiatric disorder as outcome variable. The crude model adjusted for principal components, sex, and birth-year. The adjusted model additionally adjusted for all psychiatric PRSs. ORs correspond to the multiplicative change in odds per +1 standard deviation in PRS.

ADHD, attention deficit/hyperactivity disorder; AOR, adjusted odds ratio; EstBB, Estonian Biobank; MDD, major depressive disorder; OR, odds ratio; PTSD, post-traumatic stress disorder; PRS, polygenic risk score; STR, Swedish Twin Registry.

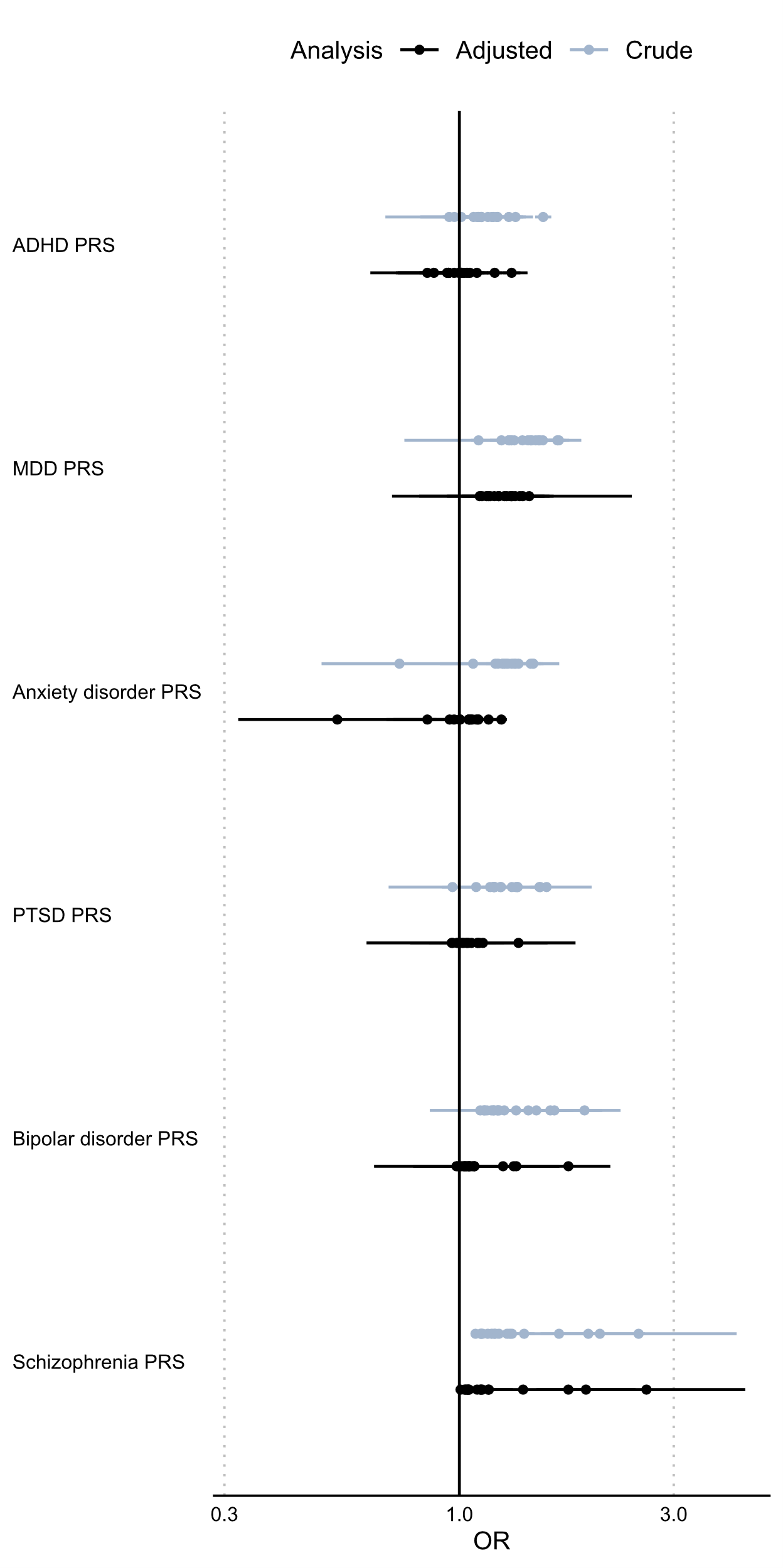

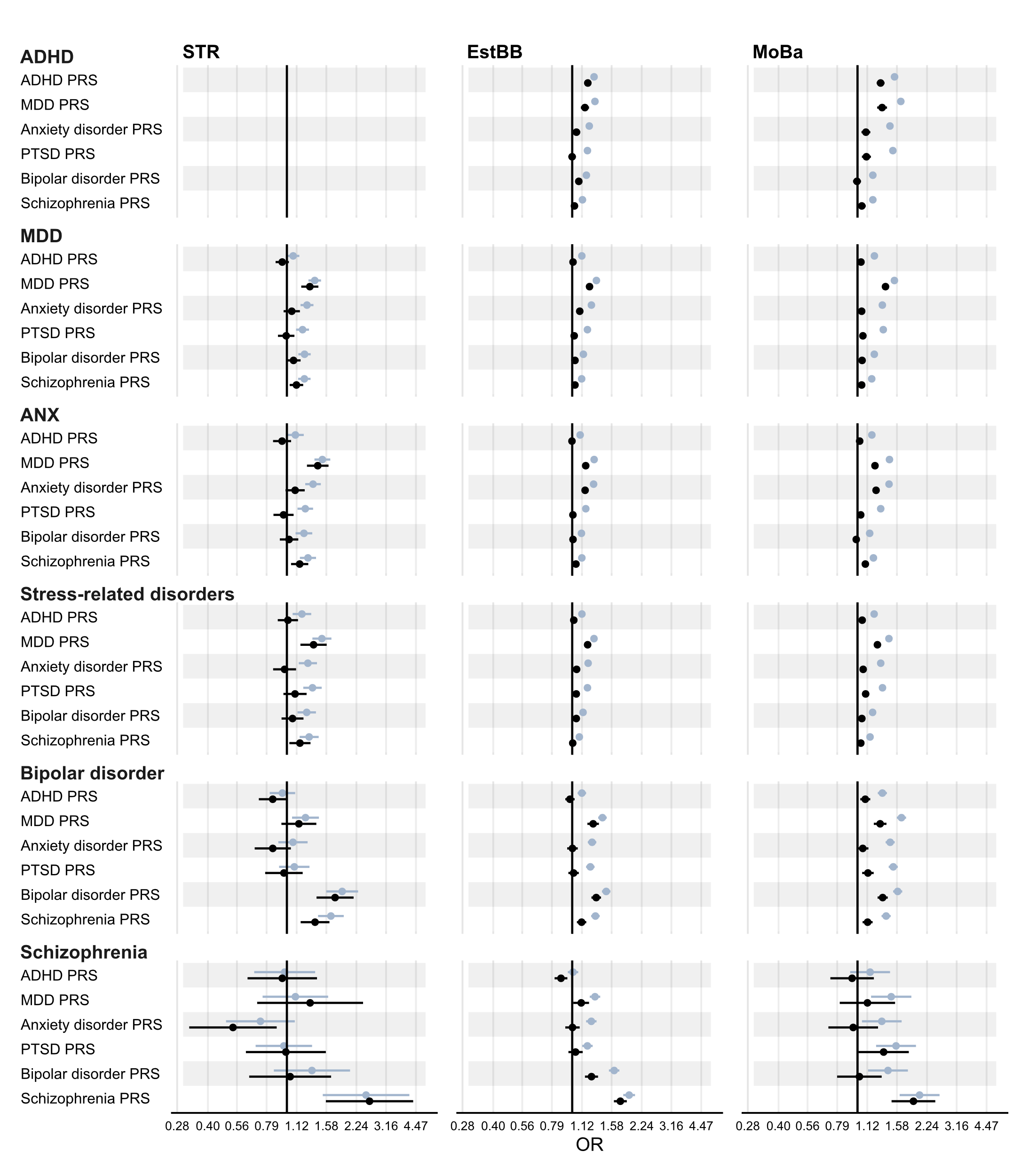

### eFigure 3. Associations of psychiatric PRSs with cardiometabolic diseases by cohort

In each cohort, ORs were computed using logistic regression models with the PRS as exposure variable and any recorded clinical diagnosis of the cardiometabolic disease as outcome variable. The crude model adjusted for principal components, sex, and birth-year. The adjusted model additionally adjusted for all other psychiatric PRSs. ORs correspond to the multiplicative change in odds per +1 standard deviation in PRS.

ADHD, attention deficit/hyperactivity disorder; AOR, adjusted odds ratio; EstBB, Estonian Biobank; MDD, major depressive disorder; OR, odds ratio; PTSD, post-traumatic stress disorder; PRS, polygenic risk score; STR, Swedish Twin Registry.

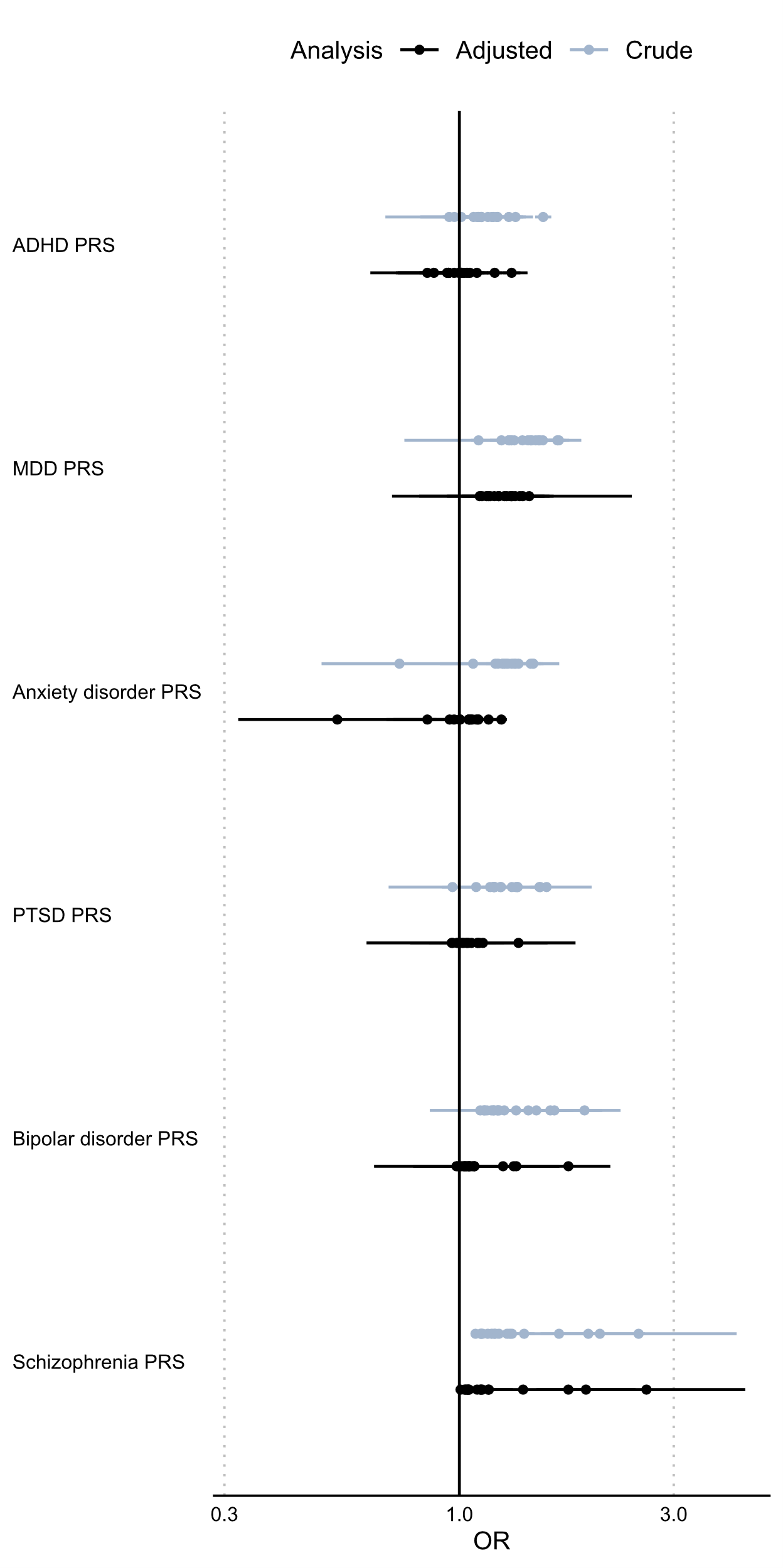

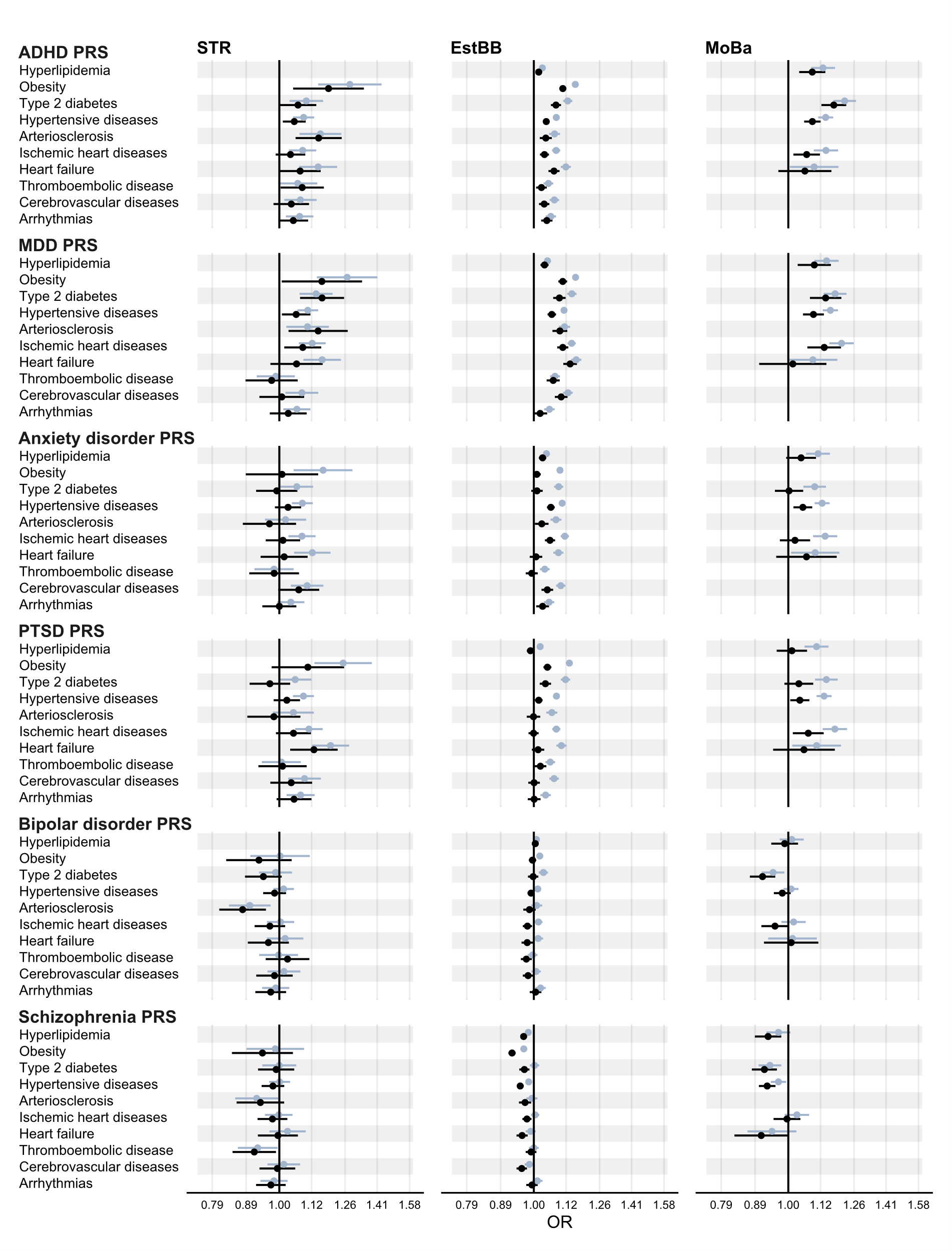

### eFigure 4. Associations of psychiatric PRSs constructed from genome-wide significant loci with cardiometabolic diseases

In each cohort, ORs were computed using logistic regression models with the PRS as exposure variable and any recorded clinical diagnosis of the cardiometabolic disease as the outcome variable. The model adjusted for principal components, sex, and birth-year. PRSs were derived using clumping and thresholding on *P*<5×10^-8^. ORs were computed from cohort-specific ORs using inverse variance weighting. ORs correspond to the multiplicative change in odds per +1 standard deviation in PRS.

ADHD, attention deficit/hyperactivity disorder; C+T, clumping and thresholding; MDD, major depressive disorder; OR, odds ratio; PTSD, post-traumatic stress disorder; PRS, polygenic risk score.

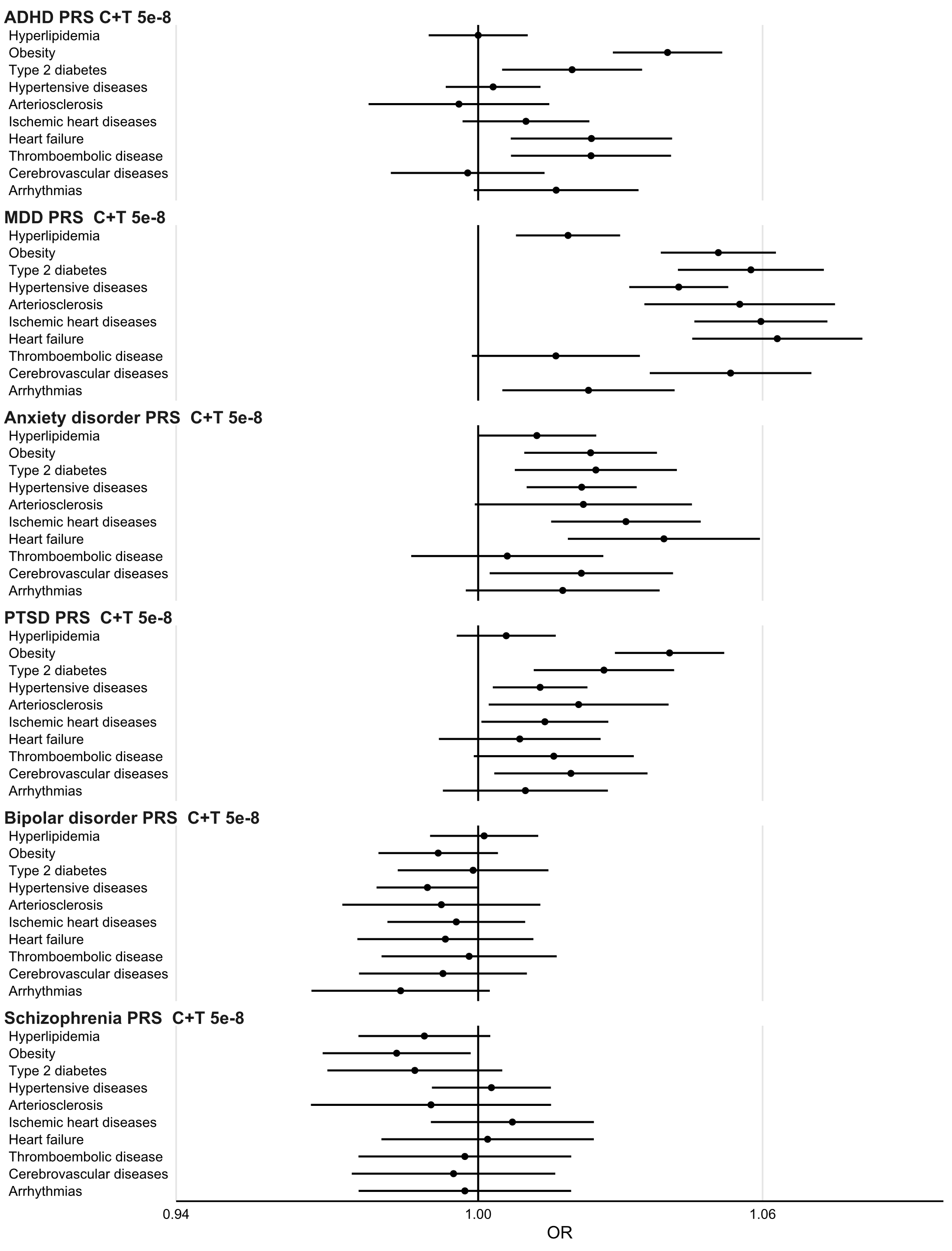

### eFigure 5. Associations of psychiatric PRSs constructed from genome-wide significant loci with cardiometabolic diseases by cohort

ORs were computed using logistic regression models with the PRS as exposure variable and any recorded clinical diagnosis of the cardiometabolic disease as the outcome variable. The model adjusted for principal components, sex, and birth-year. PRSs were derived using clumping and thresholding on *P*<5×10^-8^. ORs correspond to the multiplicative change in odds per +1 standard deviation in PRS. ADHD, attention deficit/hyperactivity disorder; EstBB, Estonian Biobank; C+T, clumping and thresholding; MDD, major depressive disorder; OR, odds ratio; PTSD, post-traumatic stress disorder; PRS, polygenic risk score; STR, Swedish Twin Registry.

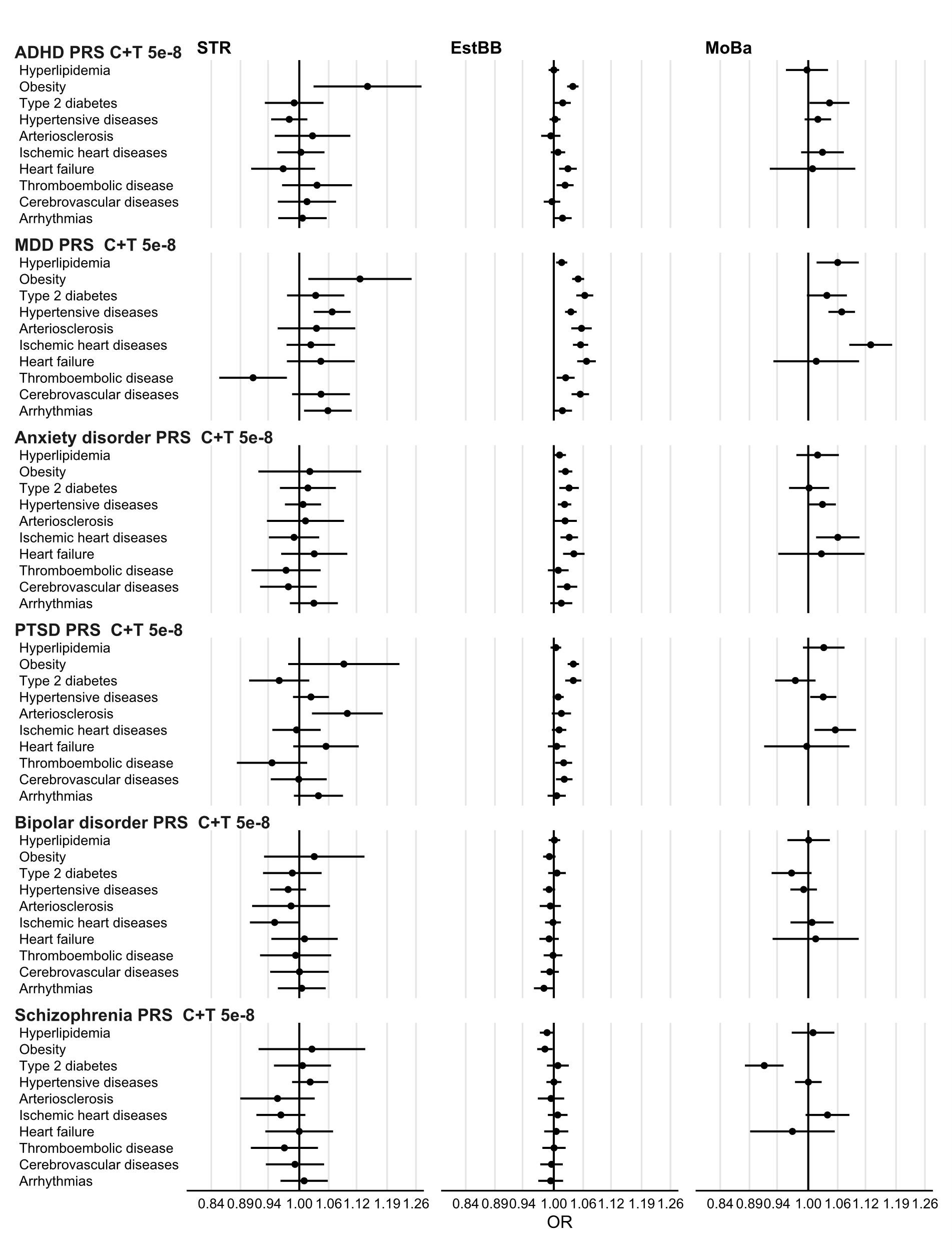

### eMethods

#### Genotyping

##### Swedish Twin Registry

For the TwinGene study, after excluding samples with DNA concentration below 20 ng/µl, 10,947 samples were processed using the Ricopili^1^ pipeline for quality control (QC).

After removing single nucleotide polymorphisms (SNPs) based on missingness > 0.05 (N = 1,709), 29 samples were removed due to any of the following: per-sample call rate < 0.98; excessive heterozygosity (FHET outside +/- 0.2); sex mismatch, and 41,972 out of 731,442 SNPs were removed due to any of the following: per-SNP call rate < 0.98; invariant; Hardy-Weinberg disequilibrium (P<1e-6). Furthermore, 47,969 SNPs with MAF < 1% were excluded, leaving 641,501 SNPs.

By projecting the first two principal components (PCs) of the study sample to the reference panel of 1000 Genome global population, we identified 6 samples as non-European ancestral outliers, whose first two PCs exceeded 6 standard deviations from the mean values of the European samples in the reference population. After excluding ancestral outliers, the same QC procedure as described above was reiterated, leading to a further exclusion of 1 sample and 4 SNPs. The dataset used for imputation thus consisted of 10,911 samples and 641,497 SNPs. Related individuals were not excluded in QC.

Of the SNPs passing QC, 638,601 were successfully aligned to the forward genomic strand and matched to the reference panel. We then used the Sanger imputation service <https://imputation.sanger.ac.uk/> to impute the post-QC genotype data to the reference panel of Haplotype Reference Consortium data^2^ (HRC1.1). EAGLE2^3^ and PBWT^4^ were used for prephasing and imputing respectively. 40,359,612 SNPs were available after the HRC imputation (before any post-imputation quality filters). After imputation, data was converted to plink2 pgen format <https://www.cog-genomics.org/plink/2.0/formats#pgen> and filtered to exclude imputed markers with imputation INFO score < 0.1, and MAF < 0.005. The final dataset included 9,113,633 markers.

We used a similar procedure for the SALTY sample. We processed 5,546 samples using the Ricopili pipeline. After QC, the dataset consisted of 5,477 samples with 300,920 SNPs, which, after imputation, addition of non-genotyped monozygotic co-twins, and further QC yielded a final dataset of 6,611 samples with 9,111,991 markers.

##### Estonian Biobank

The samples from the Estonian Biobank (EstBB), were genotyped at the Core Facility of Genomics, Institute of Genomics, University of Tartu, using the Global Screening Arrays from Illumina (GSAv1.0, GSAv2.0, GSAv2.0_EST, and GSAv3.0_EST). Genotyped samples were exported to PLINK format using GenomeStudio v2.0.4. During the quality control, all individuals with call-rate < 95% or mismatching sex that was defined based on the heterozygosity of the X chromosome and sex in the phenotype data were excluded. Variants were excluded if the call-rate was < 95% or if the Hardy-Weinberg disequilibrium p-value was <1e-4 (autosomal variants only). Variant positions were updated to genome build 37. All AT and GC variants, indels, and variants with MAF<1% were excluded. In addition, SNPs that showed potential traces of batch bias were removed. Prephasing was done using the EAGLE2 v2.4.1 software. Imputation was carried out using Beagle 5.4^5^ (beagle.22Jul22.46e). An Estonian population specific imputation reference of 2,056 WGS samples was used^6^. Further, EstBB samples were combined with the 1,000 genomes phase 3 dataset for ancestry analysis. Participants with non-European assigned group ancestry or who deviated ±3SD from the samples’ heterozygosity rate mean were removed from the analysis.

##### MoBa

We included MoBa participants that were genotyped. Blood samples were obtained from both parents during pregnancy and from mothers and children (umbilical cord) at birth^7^. Genotyping of participants has been performed in many batches as parts of several research projects. Details on genotyping procedure, imputation, and quality control are described in detail elsewhere^8^.
