## Supplement 2 for "Associations of Genetic Liability to Six Psychiatric Disorders With Cardiometabolic Diseases"

Nora I. Strom^1, 2, 3, 4^; Brad Verhulst^5^; Silviu-Alin Bacanu^6^; Rosa Cheesman^7^; Kirstin L. Purves^8^; Hüseyin Gedik^9, 10, 11^; Brittany L. Mitchell^12, 13^; Alex S. Kwong^14, 15^; Annika B. Faucon^16^; Kritika Singh^17, 18^; Sarah Medland^12^; Lucia Colodro-Conde^12, 19^; Kristi Krebs^20^; Per Hoffmann^21, 22^; Stefan Herms^21, 23, 22^; Jan Gehlen^24^; Stephan Ripke^25, 26^; Swapnil Awasthi^25^; Teemu Palviainen^27^; Elisa M. Tasanko^28^; Roseann E. Peterson^9, 6^; Daniel E. Adkins^29^; Andrey A. Shabalin^29^; Mark J. Adams^30^; Matthew H. Iveson^30^; Archie Campbell^31^; Laurent F. Thomas^32, 33, 34, 35^; Bendik S. Winsvold^36, 37, 38^; Ole Kristian Drange^39, 40, 41, 42, 43^; Sigrid Børte^44, 45, 37^; Abigail R. ter Kuile^8, 46, 47^; Joonas Naamanka^48, 49^; Tan-Hoang Nguyen^11^; Sandra M. Meier^50^; Elizabeth C. Corfield^51, 52^; Laurie Hannigan^52, 51, 53^; Daniel F. Levey^54, 55^; Darina Czamara^56^; Heike Weber^57^; Karmel W. Choi^58, 59^; Giorgio Pistis^60^; Baptiste Couvy-Duchesne^12, 61, 62^; Sandra Van der Auwera^63^; Alexander Teumer^63; 64^; Robert Karlsson^65^; Miguel Garcia-Argibay^66, 65^; Donghyung Lee^67^; Rujia Wang^68^; Ottar Bjerkeset^69, 39^; Eystein Stordal^70, 39^; Julia Bäckman^3^; Giovanni A. Salum^71, 72^; Clement C. Zai^73, 74, 75, 76, 77^; James L. Kennedy^73, 74, 75^; Gwyneth Zai^73, 74, 75^; Arun K. Tiwari^73, 74, 75^; Stefanie Heilmann-Heimbach^21^; Börge Schmidt^78^; Jaakko Kaprio^27^; Martin, M. Kennedy^79^; Joseph Boden^80^; Alexandra Havdahl^51, 53, 7, 14^; Christel M. Middeldorp^81, 82^; Fabiana L. Lopes^83, 84^; Nirmala Akula^85^; Francis J. McMahon^85, 86^; Elisabeth B. Binder^56^; Lydia Fehm^87^; Andreas Ströhle^88^; Enrique Castelao^89^; Henning Tiemeier^90, 91^; Dan J. Stein^92^; David Whiteman^93^; Catherine Olsen^93^; Zachary Fuller^94^; Xin Wang^94^; Naomi R. Wray^62, 95^; Enda M. Byrne^81^; Glyn Lewis^96^; Nicholas J. Timpson^53, 14^; Lea K. Davis^17^; Ian B. Hickie^97^; Nathan A. Gillespie^6^; Lili Milani^20^; Johannes Schumacher^24^; David P. Woldbye^98^; Andreas J. Forstner^21, 99, 24^; Markus M. Nöthen^21^; Iiris Hovatta^100^; John Horwood^80^; William E. Copeland^101^; Hermine H. Maes^11, 6, 102^; Andrew M. McIntosh^30^; Ole A. Andreassen^41, 42, 103^; John-Anker Zwart^44, 37, 45^; Ole Mors^104, 105^; Anders D. Børglum^4, 105, 106^; Preben B. Mortensen^107^; Helga Ask^51, 7^; Ted Reichborn-Kjennerud^51, 41^; Jackob M. Najman^108^; Murray B. Stein^109, 110^; Joel Gelernter^55, 111, 112^; Yuri Milaneschi^113^; Brenda W. Penninx^113^; Dorret I. Boomsma^114, 115^; Eduard Maron^116, 117^; Angelika Erhardt-Lehmann^56, 118^; Christian Rück^3^; Tilo T. Kircher^119^; Christiane A. Melzig^120, 121^; Georg W. Alpers^122^; Volker Arolt^123^; Katharina Domschke^124, 125^; Jordan W. Smoller^59, 58^; Martin Preisig^60^; Nicholas G. Martin^12^; Michelle K. Lupton^12, 13, 126^; Annemarie I. Luik^127^; Andreas Reif^128^; Hans J. Grabe ^63^; Henrik Larsson^66, 65^; Patrik K. Magnusson^65^; Albertine J. Oldehinkel^129^; Catharina A. Hartman^129^; Gerome Breen^8^; Anna R. Docherty^130, 131, 6^; Hilary Coon^130^; Rupert Conrad^132^; Kelli Lehto^20^; Jürgen Deckert^118^; Thalia C. Eley^8^; Manuel Mattheisen^133^, ^134, 2^; John M. Hettema^5^.

^1^: Department of Psychology, Humboldt-Universität zu Berlin, Berlin, Germany

^2^: Institute of Psychiatric Phenomics and Genomics (IPPG), University Hospital, LMU Munich, Munich, Germany

^3^: Centre for Psychiatry Research, Department of Clinical Neuroscience, Karolinska Institutet & Stockholm Health Care Services, Region Stockholm, Stockholm, Sweden

^4^: Department of Biomedicine, Aarhus University, Aarhus, Denmark

^5^: Psychiatry and Behavioral Sciences, Texas A&M University, College Station, Texas, USA

^6^: Psychiatry, Virginia Commonwealth University, Richmond, Virginia, USA

^7^: PROMENTA Centre, Department of Psychology, University of Oslo, Oslo, Norway

^8^: Social, Genetic and Developmental Psychiatry Centre, Institute of Psychiatry, Psychology and Neuroscience, King's College London, London, UK

^9^: Institute for Genomics in Health, Department of Psychiatry and Behavioral Sciences, State University of New York Downstate Health Sciences University, Brooklyn, New York, USA

^10^: Life Sciences, Integrative Life Sciences Doctoral Program, Virginia Commonwealth University, Richmond, Virginia, USA

^11^: Human and Molecular Genetics, Virginia Institute for Psychiatric and Behavioral Genetics, Virginia Commonwealth University, Richmond, Virginia, USA

^12^: Brain and Mental Health Program, QIMR Berghofer Medical Research Institute, Brisbane, Queensland, Australia

^13^: Faculty of Medicine, Queensland University , Brisbane, Queensland, Australia

^14^: Bristol Medical School, Population Health Sciences, MRC Integrative Epidemiology Unit, University of Bristol, Bristol, UK

^15^: Centre for Clinical Brain Sciences, Division of Psychiatry, University of Edinburgh, Edinburgh, UK

^16^: Division of Medicine, Human Genetics, Vanderbilt University, Nashville, Tennessee, USA

^17^: Division of Genetic Medicine, Vanderbilt University Medical Center, Nashville, Tennessee, USA

^18^: Vanderbilt Genetics Institute, Vanderbilt University Medical Center, Nashville, Tennessee, USA

^19^: School of Psychology, The University of Queensland, Brisbane, Queensland, Australia

^20^: Estonian Genome Centre, Institute of Genomics, University of Tartu, Tartu, Estonia

^21^: Institute of Human Genetics, University of Bonn, School of Medicine & University Hospital Bonn, Bonn, Germany

^22^: Department of Biomedicine, Human Genomics Research Group, University of Basel; University Hospital Basel, Basel, Switzerland

^23^: Institute of Medical Genetics and Pathology, Medical Faculty, University Hospital Basel, Basel, Switzerland

^24^: Center for Human Genetics, University of Marburg, Marburg, Germany

^25^: Dept. of Psychiatry and Psychotherapy, Charité - Universitätsmedizin, Berlin, Germany

^26^: Analytic and Translational Genetics Unit, Massachusetts General Hospital, Boston, Massachusetts, USA

^27^: Helsinki Institute of Life Science, Institute for Molecular Medicine Finland - FIMM, University of Helsinki, Helsinki, Finland

^28^: Faculty of Medicine, Department of Psychology and Logopedics, SleepWell Research Program, University of Helsinki, Helsinki, Finland

^29^: School of Medicine, Department of Psychiatry, University of Utah, Salt Lake City, Utah, USA

^30^: Centre for Clinical Brain Sciences, University of Edinburgh, Edinburgh, UK

^31^: College of Medicine and Veterinary Medicine, Institute of Genetics and Cancer;Centre for Genomic and Experimental Medicine, University of Edinburgh, Edinburgh, UK

^32^: Department of Clinical and Molecular Medicine, Norwegian University of Science and Technology, Trondheim, Norway

^33^: HUNT Center for Molecular and Clinical Epidemiology, Department of Public Health and Nursing, Faculty of Medicine and Health Sciences, Norwegian University of Science and Technology, Trondheim, Norway

^34^: BioCore - Bioinformatics Core Facility, Norwegian University of Science and Technology, Trondheim, Norway

^35^: Clinic of Laboratory Medicine, St. Olavs Hospital, Trondheim University Hospital, Trondheim, Norway

^36^: Division of Clinical Neuroscience, Department of Research and Innovation, Oslo University Hospital, Oslo, Norway

^37^: Department of Public Health and Nursing, HUNT Center for Molecular and Clinical Epidemiology, Norwegian University of Science and Technology, Trondheim, Norway

^38^: Department of Neurology, Oslo University Hospital, Oslo, Norway

^39^: Department of Mental Health, Norwegian University of Science and Technology, Trondheim, Norway

^40^: Division of Mental Health, St. Olavs Hospital, Trondheim University Hospital, Trondheim, Norway

^41^: NORMENT Centre, University of Oslo, Oslo, Norway

^42^: Centre of Precision Psychiatry, Division of Mental Health and Addiction, Oslo University Hospital and University of Oslo, Oslo, Norway

^43^: Department of Psychiatry, Sørlandet Hospital, Kristiansand, Norway

^44^: Division of Clinical Neuroscience, Department of Research and Innovation; Musculoskeletal Health, Oslo University Hospital, Oslo, Norway

^45^: Faculty of Medicine, Institute of Clinical Medicine, University of Oslo, Oslo, Norway

^46^: National Institute for Health and Care Research (NIHR) Maudsley Biomedical Research Centre, South London and Maudsley NHS Foundation Trust, London, UK

^47^: Department of Clinical, Educational and Health Psychology, University College London, London, United Kingdom

^48^: Hector Institute for Artificial Intelligence in Psychiatry, Central Institute of Mental Health, Mannheim, Germany

^49^: SleepWell Research Program, Faculty of Medicine, University of Helsinki, Helsinki, Finland

^50^: Psychiatry, Dalhousie University, Halifax, Nova Scotia, Canada

^51^: PsychGen Centre for Genetic Epidemiology and Mental Health, Norwegian Institute of Public Health, Oslo, Norway

^52^: Nic Waals Institute, Lovisenberg Diaconal Hospital, Oslo, Norway

^53^: Bristol Medical School, Population Health Sciences, University of Bristol, Bristol, UK

^54^: Department of Psychiatry, Division of Human Genetics, Yale University School of Medicine, New Haven, Connecticut, USA

^55^: Psychiatry, Research, Veterans Affairs Connecticut Healthcare System, West Haven, Connecticut, USA

^56^: Department of Genes and Environment, Max-Planck Institute of Psychiatry, Munich, Germany

^57^: Department of Psychiatry, Psychosomatics and Psychotherapy, University Hospital of Würzburg, Würzburg, Germany

^58^: Psychiatry, Center for Precision Psychiatry, Massachusetts General Hospital, Boston, Massachusetts, USA

^59^: Psychiatry, Psychiatric and Neurodevelopmental Genetics Unit, Center for Genomic Medicine, Massachusetts General Hospital, Boston, Massachusetts, USA

^60^: Psychiatric Epidemiology and Psychopathology Research Center, Department of Psychiatry, Lausanne University Hospital and University of Lausanne, Prilly, Switzerland

^61^: ARAMIS laboratory, Paris Brain Institute, Paris, France

^62^: Institute for Molecular Bioscience, University of Queensland, Brisbane, Queensland, Australia

^63^: Department of Psychiatry and Psychotherapy, University Medicine Greifswald, Greifswald, Germany

^64^: Institute for Community Medicine, University Medicine Greifswald, Greifswald, Germany

^65^: Department of Medical Epidemiology and Biostatistics, Karolinska Institutet, Stockholm, Sweden

^66^: School of Medical Sciences, Faculty of Medicine and Health, Örebro University, Örebro, Sweden

^67^: Department of Statistics, Miami University, Oxford, Ohio, USA

^68^: Social, Genetic, and Developmental Psychiatry Centre, Institute of Psychiatry, Psychology and Neuroscience, King's College London, London, UK

^69^: Faculty of Nursing and Health Science, Nord University, Levanger, Norway

^70^: Department of Psychiatry, Hospital Namsos, Nord-Trøndelag Health Trustt, Namsos, Norway

^71^: Department of Psychiatry, Universidade Federal do Rio Grande do Sul, Porto Alegre, Rio Grande do Sul, Brazil

^72^: Child Psychiatry, National Institute of Developmental Psychiatry, São Paulo, Brazil

^73^: Tanenbaum Centre for Pharmacogenetics, Molecular Brain Sciences Department, Campbell Family Mental Health Institute, Centre for Addiction and Mental Health, Toronto, Ontario, Canada

^74^: Department of Psychiatry, Division of Neurosciences and Clinical Translation, University of Toronto, Toronto, Ontario, Canada

^75^: Institute of Medical Science, University of Toronto, Toronto, Ontario, Canada

^76^: Laboratory Medicine and Pathobiology, University of Toronto, Toronto, Ontario, Canada

^77^: Stanley Center for Psychiatric Research, Broad Institute of Harvard and MIT, Cambridge, MA, USA

^78^: Institute for Medical Informatics, Biometry and Epidemiology, University Hospital of Essen, University of Duisburg-Essen, Essen, Germany

^79^: Pathology and Biomedical Science, University of Otago, Christchurch, New Zealand

^80^: Psychological Medicine, University of Otago, Christchurch, New Zealand

^81^: Child Health Research Centre, University of Queensland, Brisbane, Queensland, Australia

^82^: Child and Youth Mental Health Service, Children's Health Queensland Hospital and Health Service, Brisbane, Queensland, Australia

^83^: National Institute of Mental Health, Human Genetics Branch, National Institutes of Health, Bethesda, Maryland, USA

^84^: Department of Psychiatry and Human Behavior, Alpert Medical School of Brown University, Providence, Rhode Island, USA

^85^: National Institute of Mental Health, Genetic Basis of Mood and Anxiety Disorders, National Institutes of Health, Bethesda, Maryland, USA

^86^: Psychiatry & Behavioral Sciences, Johns Hopkins University, Baltimore, Maryland, USA

^87^: Department of Psychology, Zentrum für Psychotherapie, Humboldt-Universität zu Berlin, Berlin, Germany

^88^: Department of Psychiatry and Psychotherapy, Campus Charité Mitte, Charité - Universitätsmedizin Berlin, Corporate member of Freie Universität Berlin and Humboldt-Universität zu Berlin, Berlin, Germany

^89^: Psychiatric Epidemiology and Psychopathology Research Center, Department of Psychiatry, Lausanne University Hospital and University of Lausanne, Prilly, Switzerland

^90^: Social and Behavioral Science, T.H. Chan School of Public Health, Harvard University, Boston, Massachusetts, USA

^91^: Child and Adolescent Psychiatry, Erasmus University Medical Center, Rotterdam, Netherlands

^92^: SAMRC Unit on Risk & Resilience in Mental Disorders, Department of Psychiatry & Neuroscience Institute, University of Cape Town, Cape Town, South Africa

^93^: Population Health Program, QIMR Berghofer Medical Research Institute, Brisbane, Australia

^94^: ^23^andMe, Sunnyvale, CA, USA

^95^: Department of Psychiatry, University of Oxford, Oxford, UK

^96^: UCL Division of Psychiatry, University College London, London, UK

^97^: Brain and Mind Centre, University of Sydney, Sydney, Australia

^98^: Department of Neuroscience, Laboratory of Neural Plasticity, University of Copenhagen, Copenhagen, Denmark

^99^: Institute of Neuroscience and Medicine (INM-^1^), Research Center Jülich, Jülich, Germany

^100^: Faculty of Medicine, Department of Psychology and Logopedics and SleepWell Research Program, University of Helsinki, Helsinki, Finland

^101^: UVM Medical Center, Department of Psychiatry, University of Vermont, Burlington, Vermont, USA

^102^: Massey Cancer Center, Virginia Commonwealth University, Richmond, Virginia, USA

^103^: K. G. Jebsen Center for Neurodevelopmental disorders, University of Oslo, Oslo, Norway

^104^: Department of Psychiatry, Psychosis Research Unit, Aarhus University Hospital, Aarhus, Denmark

^105^: The Lundbeck Foundation Initiative for Integrative Psychiatric Research, iPSYCH, Aarhus University, Aarhus, Denmark

^106^: Center for Genomics and Personalised Medicine, Aarhus University, Aarhus, Denmark

^107^: The National Centre for Register-based Research, Aarhus University, Aarhus, Denmark

^108^: Faculty of Medicine, School of Public Health, University of Queensland, Herston, Queensland, Australia

^109^: Psychiatry, University of California San Diego, La Jolla, CA, USA

^110^: School of Public Health, University of California San Diego, La Jolla, CA, USA

^111^: Psychiatry Research, Veterans Affairs Connecticut Healthcare System, West Haven, Connecticut, USA

^112^: Departments of Genetics and Neuroscience, Yale University of Medicine, New Haven, Connecticut, USA

^113^: Amsterdam Neuroscience; Amsterdam Public Health, Amsterdam University Medical Center, Amsterdam, Netherlands

^114^: Twin Register and Department of Complex Trait Genetics, Center for Neurogenomics and Cognitive Research, Vrije Universiteit Amsterdam, Amsterdam, Netherlands

^115^: Amsterdam Public Health, Amsterdam University Medical Center, Amsterdam, Netherlands

^116^: Centre for Digital Health: Department of Health Technologies, Tallinn University of Technology, Tallinn, Estonia

^117^: Department of Medicine, Centre for Neuropsychopharmacology,, Division of Brain Sciences, Imperial College London, London, UK

^118^: Department of Psychiatry, Psychosomatics and Psychotherapy, Center of Mental Health, University Hospital Würzburg, Würzburg, Germany

^119^: Department of Psychiatry, University of Marburg, Marburg, Germany

^120^: Psychology, Clinical Psychology, Experimental Psychopathology and Psychotherapy, University of Marburg, Marburg, Germany

^121^: Psychology, Biological and Clinical Psychology, University of Greifswald, Greifswald, Germany

^122^: School of Social Sciences, Department of Psychology, University of Mannheim, Mannheim, Germany

^123^: Department of Mental Health, Institute for Translational Psychiatry, University of Muenster, Muenster, Germany

^124^: Department of Psychiatry and Psychotherapy, Medical Center, Faculty of Medicine, University of Freiburg, Freiburg, Germany

^125^: German Center for Mental Health (DZPG), Partner Site Berlin, Berlin, Germany

^126^: Faculty of Health, Queensland University of technology, Queensland, Australia

^127^: Epidemiology, Erasmus University Medical Center, Rotterdam, Netherlands

^128^: Department of Psychiatry, Psychosomatic Medicine and Psychotherapy, University Hospital Frankfurt - Goethe University, Frankfurt, Germany

^129^: Psychiatry, Interdisciplinary Center Psychopathology and Emotion Regulation, University of Groningen, University Medical Center Groningen, Groningen, Netherlands

^130^: School of Medicine, Psychiatry, University of Utah, Salt Lake City, Utah, USA

^131^: School of Medicine, Psychiatry, Huntsman Mental Health Institute, University of Utah, Salt Lake City, Utah, USA

^132^: Department of Psychosomatic Medicine and Psychotherapy, University Hospital Münster, Münster, Germany

^133^: Community Health and Epidemiology, Dalhousie University, Halifax, Nova Scotia, Canada

^134^: Computer Science, Dalhousie University, Halifax, Nova Scotia, Canada

### Estonian Biobank Research Group

Andres Metspalu^1^; Lili Milani^1^; Tõnu Esko^1^; Reedik Mägi^1^; Mait Metspalu^1^; Mari Nelis^1^; Georgi Hudjashov^1^.

^1^: Estonian Genome Centre, Institute of Genomics, University of Tartu
